# State switching and high-order spatiotemporal organization of dynamic Functional Connectivity are disrupted by Alzheimer’s Disease

**DOI:** 10.1101/2023.02.19.23285768

**Authors:** Lucas Arbabyazd, Spase Petkoski, Michael Breakspear, Ana Solodkin, Demian Battaglia, Viktor Jirsa

## Abstract

Spontaneous activity during the resting state, tracked by BOLD fMRI imaging, or shortly rsfMRI, gives rise to brain-wide dynamic patterns of inter-regional correlations, whose structured flexibility relates to cognitive performance. Here we analyze resting state dynamic Functional Connectivity (dFC) in a cohort of older adults, including amnesic Mild Cognitive Impairment (aMCI, *N* = 34) and Alzheimer’s Disease (AD, *N* = 13) patients, as well as normal control (NC, *N* = 16) and cognitively “super-normal” (SN, *N* = 10) subjects. Using complementary state-based and state-free approaches, we find that resting state fluctuations of different functional links are not independent but are constrained by high-order correlations between triplets or quadruplets of functionally connected regions. When contrasting patients with healthy subjects, we find that dFC between cingulate and other limbic regions is increasingly bursty and intermittent when ranking the four groups from SNC to NC, aMCI and AD. Furthermore, regions *affected at early stages of AD pathology* are less involved in higher-order interactions in patient than in control groups, while pairwise interactions are not significantly reduced. Our analyses thus suggest that the spatiotemporal complexity of dFC organization is precociously degraded in AD and provides a richer window into the underlying neurobiology than time-averaged FC connections.

**Author Summary:** Brain functions emerge from the coordinated dynamics of many brain regions. Dynamic Functional Connectivity (dFC) analyses are a key tool to describe such dynamic complexity and have been shown to be good predictors of cognitive performance. This is particularly true in the case of Alzheimer’s Disease (AD) in which an impoverished dFC could indicate compromised functional reserve due to the detrimental effects of neurodegeneration. Here we observe that in healthy ageing dFC is indeed spatiotemporally organized, as reflected by high-order correlations between multiple regions. However, in people with aMCI or AD, dFC becomes less “entangled”, more random-like, and intermittently bursty. We speculate that this degraded spatiotemporal coordination may reflect dysfunctional information processing, thus ultimately leading to worsening of cognitive deficits.

## Introduction

Alzheimer’s Disease (AD) is the most common neurodegenerative illness with an estimated prevalence of 10-30% in people older than 65 years (Hou et al., 2019; Masters et al., 2015). Yet, despite substantial research, we are far from fully understanding the *mechanisms* that link pathophysiology to cognitive impairments. Neurodegeneration in AD has been traditionally associated with the extracellular accumulation of insoluble amyloid-β_42_ (Aβ) neuritic plaques (Glenner and Wong, 1984; Lemere et al., 1996) along with the intracellular accumulation of abnormally phosphorylated tau (pTau), that constitute the neurofibrillary tangles (Spires-Jones and Hyman, 2014). These processes yield to widespread neuronal death, synaptic loss, and atrophy (Bateman et al., 2012), with a progression of structural damages not occurring uniformly throughout the brain (Braak and Braak, 1991). However, the progression of neurodegenerative processes does not correlate linearly with the severity of cognitive impairment possibly due to a “cognitive reserve” accrued through education, cognitive training and a healthy lifestyle (Rentz et al., 2010; Snowdon, 2003). Furthermore, the severity of cognitive impairment symptoms in a patient can fluctuate substantially within the same day, faster than the time scales of neurodegeneration (Palop et al., 2006). Together, these findings suggest that AD involve alterations of neural dynamics and that these dynamical changes may be the mechanistic substrate leading to functional impairment or preservation.

As molecular and structural changes alone do not fully account for cognitive impairment, alternative studies based on Functional Connectivity (FC) analyses have sought to fill the gap. In particular, resting state FC (Fox and Raichle, 2007) quantifies brain-wide correlations of BOLD signals, capturing interactions between regions. In this context it has been suggested that structural alterations in AD lead to FC changes (Dennis and Thompson, 2014), and that the early manifestation of Aβ toxicity preceding overt atrophy can be detected using resting state functional Magnetic Resonance Imaging (rsfMRI) (Hedden *et al*., 2009; Sheline *et al*., 2010*a*; Sheline *et al*., 2010*b*; Mormino *et al*., 2011). Changes in FC in AD include reduced connectivity within the default mode network (DMN, Greicius *et al*., 2004; Rombouts *et al*., 2005; Wang *et al*., 2006, 2007; Sorg *et al*., 2007; Fleisher *et al*., 2009; Zhang *et al*., 2009, 2010; Jones *et al*., 2011; Petrella *et al*., 2011), in a spatially non-uniform fashion (Damoiseaux et al., 2012). Besides Aβ, the deposition of pTau affects FC as well (Franzmeier et al., 2022). Furthermore, additional FC alterations have been reported, leading to functional disconnection between hemispheres (Shi et al., 2020; Wang et al., 2015) and a reduction of small-world topology (Brier et al., 2014; Sanz-Arigita et al., 2010; Stam et al., 2009, 2007; Supekar et al., 2008).

More recently, investigations of FC in AD have been extended to encompass time-varying, rather than time-averaged FC. Indeed, rsfMRI networks undergo a continuous reconfiguration of their weighed topology, and the statistical structure of spontaneous network reconfiguration carries information potentially useful to discriminate cohorts (Calhoun et al., 2014; Hutchison et al., 2013; Preti et al., 2017). The flexibility of dynamic Functional Connectivity (dFC) has been shown to correlate with cognitive performance (Bassett et al., 2011; Battaglia et al., 2020; Braun et al., 2015; Jia et al., 2014; Lombardo et al., 2020; Shine et al., 2016). In this view, ongoing variability of FC networks is not noise but rather, an actual resource subserving computation. The capacity to actively maintain a spatiotemporally organized yet variable dFC would confer the system resilience to cope with variable cognitive and environmental conditions (Lombardo et al., 2020). Hence, the preservation of a “healthy” structured dFC variability may provide a form of functional compensation and a likely neural substrate for “cognitive reserve” (cf. also other studies linking mental training with enhanced dFC variability, e.g. Premi et al., 2020). Conversely, dynamic FC-based metrics thus promise to better characterize the impact of AD pathology.

A number of studies have quantified dFC changes in healthy aging (Battaglia et al., 2020; Davison et al., 2016; Hutchison and Morton, 2015; Lavanga et al., 2022; Petkoski et al., 2023; Qin et al., 2015; Viviano et al., 2017) and in conditions such as schizophrenia (Damaraju et al., 2014; Sakoğlu et al., 2010), epilepsy (Liao et al., 2014; Liu et al., 2017) and Parkinson’s disease (Fiorenzato et al., 2019; Kim et al., 2017). In AD, probabilities of temporal transitions between alternative FC states have been shown to be altered (Jones *et al*., 2011; Fu et al., 2019; Gu et al., 2020; Schumacher et al., 2019). Moreover, machine learning applications have achieved greater accuracy in differentiating between healthy control and aMCI or AD subjects when trained with dFC-based rather than static FC metrics (Chen et al., 2017, 2016; de Vos et al., 2018; Wee et al., 2016). Although the contributions of these studies are promising, they are largely descriptive and do not propose an explicit theory of why dFC changes lead to functional consequences. Furthermore, the plethora of methods for dFC quantification (Hutchison et al., 2013; Preti et al., 2017) – from extracting discrete FC states (Allen et al., 2014; Thompson and Fransson, 2016) to continuously time-resolved approaches (Battaglia et al., 2020; Lindquist et al., 2014)– hinder the convergence of results.

Here, we start from a theoretical tenet: efficient cognition requires spatiotemporally organized FC variability, which is neither trivial, nor random, but complex. This assumption is based on empirical evidence. Fluctuations in dFC are not a mere unstructured “Drunkard’s walk”: More highly structured dFC trajectories are observed in individuals with higher performance on general cognition domains (Battaglia et al., 2020; Lavanga et al., 2022). Further, individual FC links do not fluctuate independently but with network reconfigurations governed by higher order coordination patterns, manifest by: non-trivial inter-link covariance patterns (Davison et al., 2015; Faskowitz et al., 2020; Petkoski et al., 2023); “back-bones” partially scaffolding dFC (Braun et al., 2015); and dFC flowing under the influence of competing “meta-hubs (Lombardo et al., 2020). Reiterating, our hypothesis suggests that spatiotemporal structure of dFC between order and randomness allows for rich *computation* to emerge from the systems’ collective activity (cf. Crutchfield, 2012). Correspondingly, we predict that individuals with higher cognitive performance should display an enhanced organization of dFC compared to those with impaired cognition (aMCI or AD) in which, conversely, a loss of dFC spatiotemporal organization should be evident.

Here we analyze resting-state fMRI data acquired from individuals with better-than-normal or normal cognitive performance –“supernormal” (SNC) and “normal controls (NC)– and those clinically diagnosed with amnestic Mild Cognitive Impairment (aMCI) or Alzheimer’s Disease (AD). We first characterized dFC across groups using two complementary methods. First, we use a *state-based dFC analysis* paradigm, in which we assume the existence of a small set of possible discrete FC configurations and quantify dwell times in different states and the temporal stability of different FC network links along state switching transitions (Thompson and Fransson (2016)). Second, we use a *state-free dFC analysis* paradigm, where FC networks are described as continually morphing in time. Through these complementary but convergent approaches, as described in the following, we find that the fluctuations of different links show different degrees of mutual inter-dependence across the considered groups, shifting from a “liquid-like” dFC (flexible but constrained) for SNC and NC toward a “gas-like” dFC (uncorrelated and disordered) for patient groups. We also show that these changes in dFC coordination cannot be fully accounted by changes occurring at the level of ordinary pairwise FC, but stem from the weakening of genuine higher-order interactions observed especially for regions which are among the first to be physio-pathologically affected by AD.

## Results

### FC and dFC across a spectrum of cognitive performance

We considered an fMRI dataset including resting state sessions from subjects with varying degrees of cognitive skills. As our interest focusses not only on disease but also in healthy cognition, healthy controls were subclassified in two groups (SNC and NC) based primarily on composite memory Z scores to define the SNC and NC groups. That is, SNC had a higher performance in the composite memory scores (Z > 1.5) and at least a Z > 0.7 in all other cognitive domains (attention, language, visuo-spatial and executive; see *Materials and Methods* for more details). Healthy control subjects between NC and SNC or below NC were not considered in the study. As shown in Fig. 1A, from 73 subjects, 10 were classified as *supernormal controls* (SNC), 16 as *normal controls* (NC), 34 as amnesic *mild cognitive impairment* (aMCI), and 13 as *Alzheimer’s disease* (AD). Across the four clinical groups, there were no significant differences in age and sex.

**Fig. 1.**
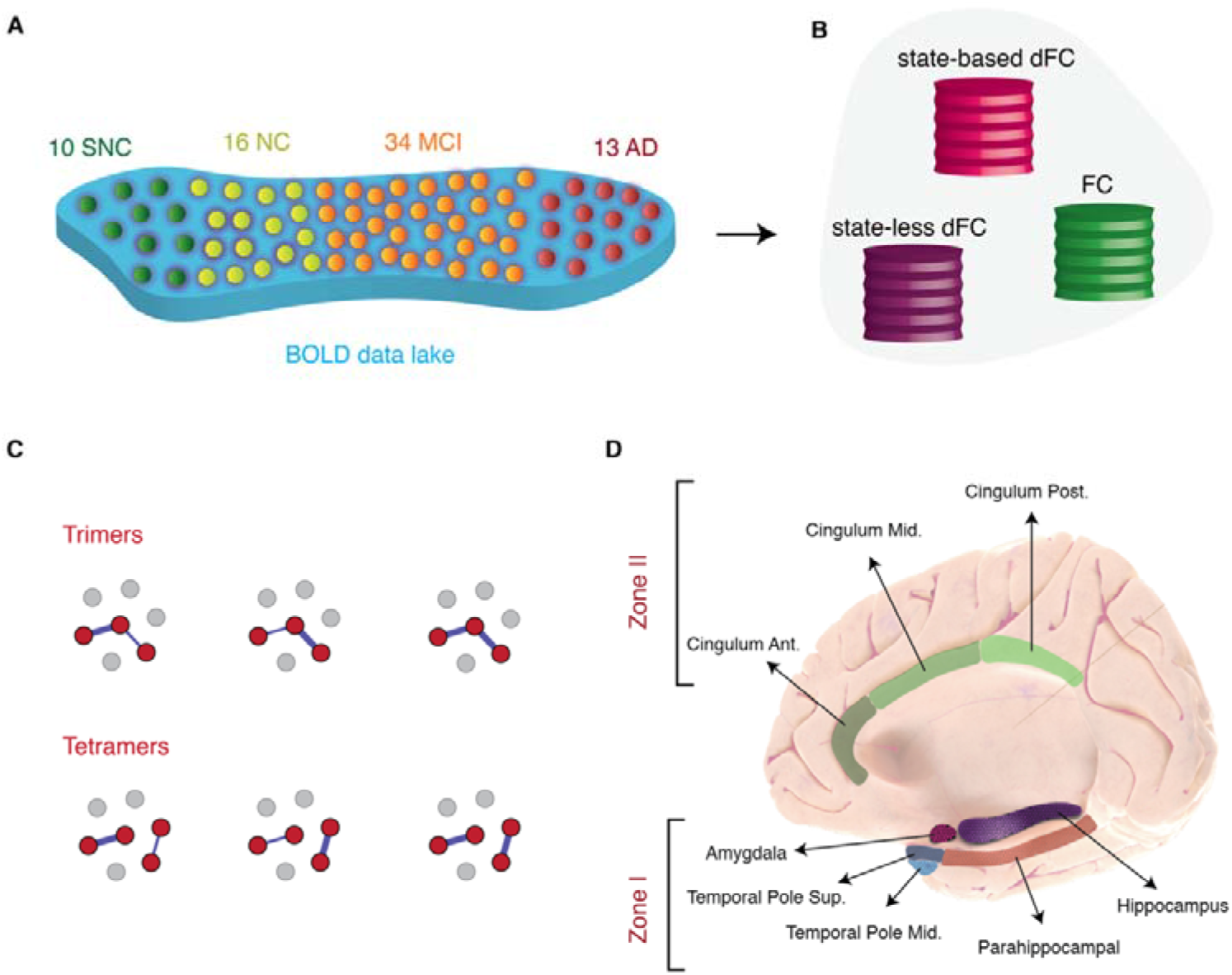
Overview of approaches. (**A**) Subjects were stratified in 4 different clinical groups: Supernormal controls (SNC), Normal controls (NC), amnesic MCI (aMCI) and Alzheimer’s disease (AD) (**B**) We used two dynamic functional connectivity (dFC) methods to study the spatiotemporal properties of resting-state fMRI signals: A state-based dFC called point-based method (PBM) and a state-free dFC method called meta-connectivity (MC) approach. Both approaches address the dynamics of pairwise links of interactions, which we call here “dimers”. (**C**) The study of coordinated fluctuations of dimers is at the core of the MC approach. Coordination can occur between dimers converging on a common root (“trimers”) or between non-incident dimers (“tetramers”). (**D**) We focused on a limbic subnetwork based on the AAL parcellation that was divided into two zones: a ventrally located “Zone I” that included the temporal pole (superior and medial), parahippocampal gyrus, hippocampus proper and amygdala; and a dorsally located Zone II included the anterior, medial and posterior cingulate cortices.

Based on rsfMRI time-series from these cohorts, we then computed (and compared across groups) a variety of static and dynamic Functional Connectivity (FC and dFC) metrics, extracted with complementary approaches, assuming or not the existence of discrete FC states in time (Fig. 1B). Importantly, as detailed below, we did not uniquely consider pairwise interactions between two brain regions at a time, but also considered more complex coordination patterns between larger groups of regions. Classic FC links express the existence of a correlation between the BOLD fluctuations of two brain regions and are represented as a link between two regional nodes: we refer hence to them as *dimers*, since they are computed out of two parts. In classical FC analyses, dimers are static, as their strength is averaged over the duration of complete resting state sessions. In dFC analyses, however dimer strengths fluctuate in time. We can thus also compute correlations between different dimers. Estimating these “correlations between correlations” requires jointly monitoring the BOLD fluctuations of three (Fig. 1C, top) or four (Fig. 1C, bottom) regions, hence the names of *trimers* and *tetramers* –collections of three or four parts, respectively– used in the following.

We chose to focus in this study on dFC within a network of limbic brain regions of particular interest (Fig. 1D). The rationale was twofold: first, the regions included in the chosen limbic subnetwork are highly interconnected brain regions that degenerate early in the disease process (Arnold et al., 1991; Braak and Braak, 1991); second, previous modelling work confirmed their central role in shaping the evolution of FC alterations comparing healthy controls to aMCI or AD stages (J. Zimmermann et al., 2018).

### State-based dFC: two zones and four dFC states

In order to assess FC changes along time, we started with a state-based dFC approach, called the point-based method (PBM) and first introduced by Thompson and Fransson (2016). In this framework, different instantaneous images of brain-wide BOLD activation are first clustered via an unsupervised procedure into *K* states, and state-specific FC matrices FC^(λ)^ are constructed by evaluating BOLD correlations limited to timeframes assigned to a given state cluster (λ = 1…*K*, see *Materials and Methods* for details). Fig. 2A show the weighed adjacency matrices FC^(λ)^ (obtained as centroids of their respective cluster) for each of four different states of dFC, called *S-graphlets* by Thompson and Fransson (2016). An alternative graph representation of these templates is shown in Fig. S1A. The optimal number of *K* = 4 was determined based on a statistical elbow criterion (Fig. S1B) and confirmed post-hoc by the consistency of our results.

**Fig. 2.**
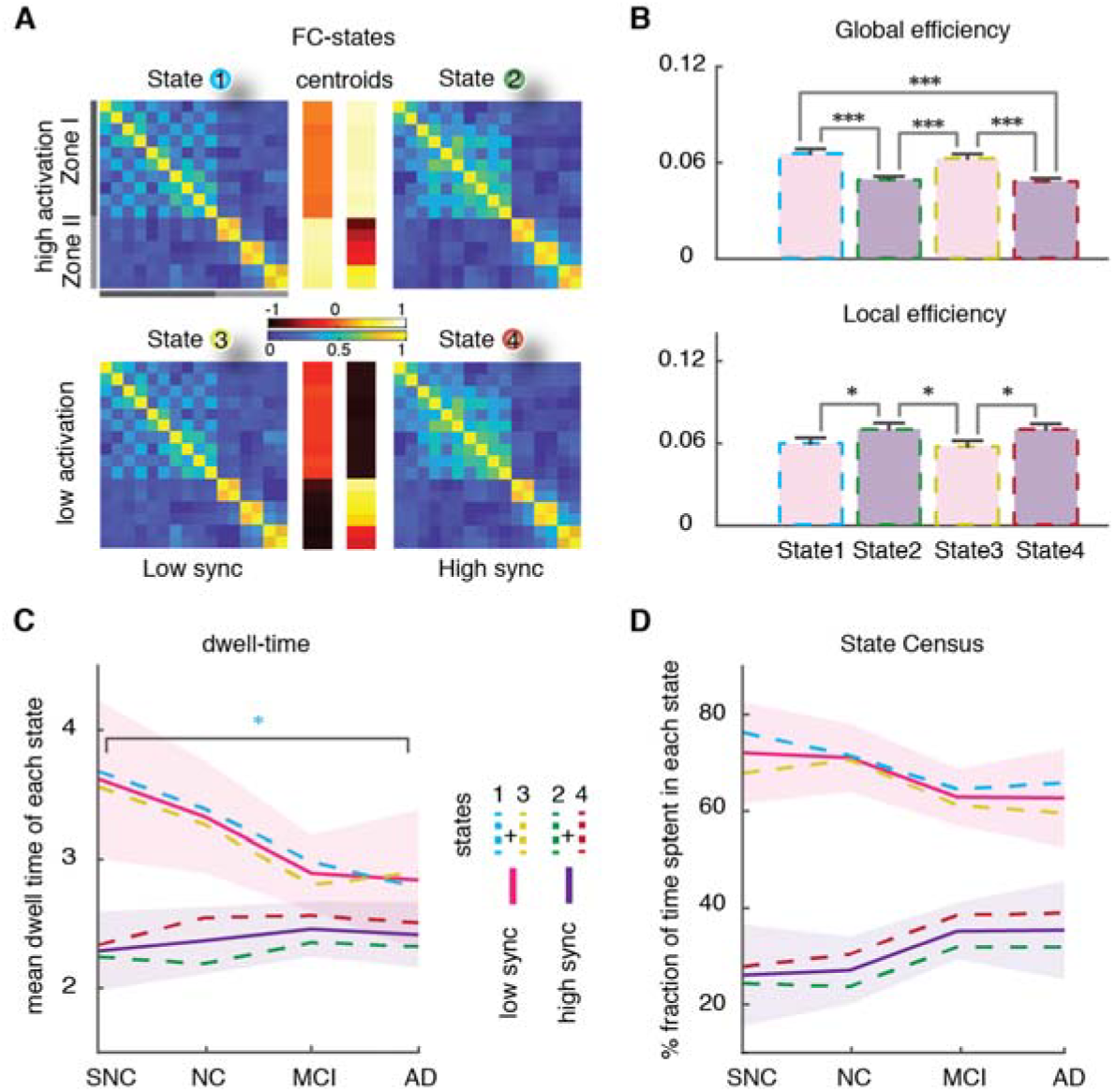
State-based dynamic Functional Connectivity (dFC) analyses: four dFC states. **(A)** BOLD time-series of all subjects were concatenated temporally and then z-scored and clustered based on BOLD activation to extract four states. The associated FC-state matrices (FC^(λ)^, λ = 1…4) were constructed by evaluation BOLD fluctuation correlations limited to time-points within a given state (cf. also Fig. S1A). The centroids of activation of four states (middle) distinguished two subsets of regions (*Zone I* and *Zone II*) where their activity was transiently higher or lower than average. States 1 and 2 (or 3 and 4) showed *above (*or *below) average level activation* for zones II and I, respectively, therefore were labelled as *high (*or *low) activation* states. We referred to states 2 and 4 as *high synchronization* states because the FC connection weights within zone I tended to be stronger than states 1 and 3 (*low synchronization*; average within zone I FC weights = 0.23 ± 0.16 for states 1 and 3 vs = 0.29 ± 0.18 for states 2 and 4). (**B**) Global and Local efficiency as measure of robustness in the communication pathways can be established between regions and was applied on the FC-states. States 1 and 3 with low synchronization showed higher global and lower local efficiency compared to high synchronization states 2 and 4. (**C**) States with low synchronization showed decrease in mean dwell-time across clinical groups (∼3.6 TR = 7.4 s, for SNC; ∼2.8 TR = 5.7 s, for AD), where the decrease of state 1 was significant (blue; p-value ∼ 0.032; Mann-Whitney U test). States 2 and 4 showed a slight increase from the control groups to the patient groups. A decrease in average dwell-time of states with relatively higher global efficiency indicates that they are less stable. (**D**) Analogously, the relative fraction of time spent in states with low synchronization was decreased in aMCI and AD compared to NC. Note the increase from SNC to AD groups for states with high synchronization.

Based on these four dFC states, we obtained the spatial profile of neural activation across regions (Fig. 2A). The spatial organization of the observed neural activation profiles naturally suggests, in this study, to group the regions in two subsets, characterized by having an activity level transiently higher or lower than their average level. We defined *“zone I”* as the subset of ventral limbic regions including amygdala, temporal pole (superior and medial), hippocampus, and parahippocampal gyrus. *“Zone II”*, included the cingulate gyrus (anterior, medial, and posterior). In states 1 and 2, zone II (dorsal regions) and zone I (ventral regions) were respectively active *above* average level (high activation states). In contrast, in states 3 and 4, zone II and zone I regions were respectively active *below* average levels (low activation states).

Furthermore, these four states were noted based on the topology of their FC^(λ)^ networks and the level of internal synchronization within zone I. Quantitatively, connection weights between regions within zone I tended to be stronger for states 2 and 4 than for states 1 and 3 (average within zone I FC weights = 0.23 ± 0.16 for states 1 and 3 vs = 0.29 ± 0.18 for states 2 and 4). Hence, states 2 and 4 displayed higher internal synchrony, in contrast to states 1 and 3. Then we computed local and global efficiency metrics (Achard and Bullmore, 2007; Latora and Marchiori, 2001) for the four FC^(λ)^ networks. Global efficiency quantifies how well communication pathways can be established between any two nodes in a weighed network. Local efficiency quantifies the robustness of communication and the possibility to find alternative routes if local connectivity is disrupted. We found that the high sync states 2 and 4 have a lower global efficiency (Fig. 2B; Mann-Whitney U test, p < 0.001) but a greater local efficiency (Fig. 2B, Mann-Whitney U test, p ∼ 0.023), reflecting a denser within-zone but a weakened between-zone connectivity (average between zone I and zone II FC weights = 0.026 ± 0.069 for states 1 and 3 vs = -0.013 ± 0.071 for states 2 and 4).

Thus, in short, the overall four states that we find are obtained as combinations of two qualitatively different network topologies an two possible levels of activation, so that each topology can exist in a low and high activity versions.

### Stability of globally efficient dFC states decreases along the clinical spectrum

We quantified the stability of dFC both by the longer or shorter duration of transient epochs within a given state (average *dwell time*, Fig. 2C) and by the overall time fraction spent within a state (average *state census*, Fig. 2D). As shown in Fig. 2C, group differences were identified in the mean dwell-time of low sync states, with longer dwell-time for the two control groups (∼3.6 TR = 7.4 s, for SNC at one extreme) and shorter for the MCI and AD groups (∼2.8 TR = 5.7 s, for AD at the other extreme). However, the mean dwell-time of high sync states were not different.

Analogously, Fig. 2D shows that the relative fraction of time spent in low sync states decreased in aMCI and AD compared to healthy controls (ranging from 62% for AD to 72% for SNC).

In summary, low-sync and globally efficient dFC states were less frequent and more transient in aMCI and AD, suggesting a reduction of their overall stability.

### Inter-zone dFC dimers are more intermittent in patient than in control groups

The next step, also following Thompson and Fransson (2016), was to map a state-based dFC temporal network to each subject’s resting-state acquisition. To do so, we constructed a sequence of network time-frames FC*(t)* set to be equal to the FC^(λ)^ graph specific for the state λ visited at time *t* (Fig. 3A; see *Materials and Methods* for details). Thompson and Fransson (2016) called such a temporal network a *T-graphlet*.

**Fig. 3.**
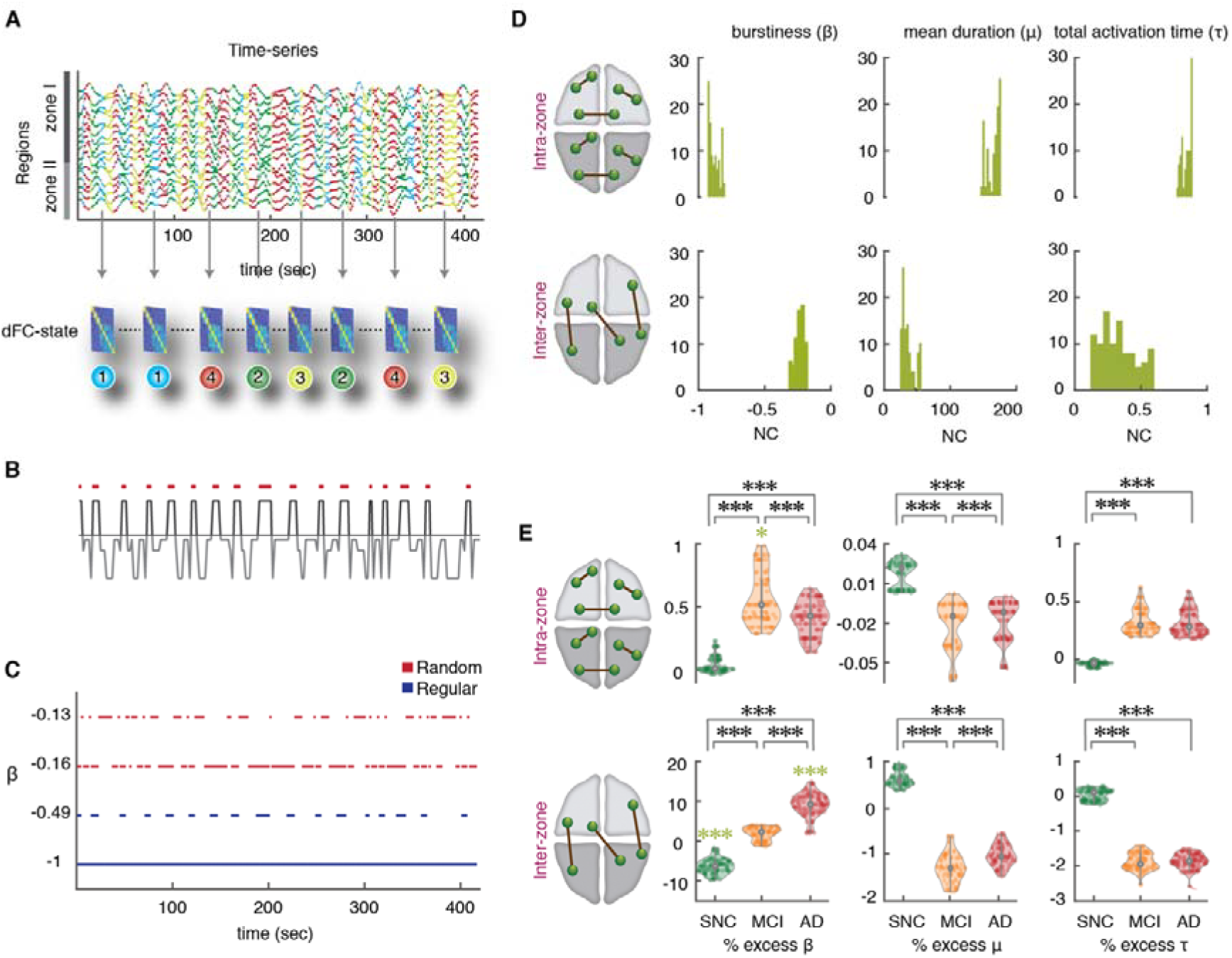
State-based dFC analyses: increase of intermittency in inter-zone links. (**A**) To construct the state-based dFC temporal network, a specific FC^(λ)^ graph was assigned to each BOLD signal intensity time-point (we show here 416 time-points = 20 minutes of rsfMRI acquisition, for two concatenated subjects). Consequently, there is a time-course for every FC links where they can assume up to four possible different strength values (link dynamics due to state switching). (**B**) The temporal organization of link fluctuations can be assessed by determining intervals of link activation and inactivation (via a thresholding of dynamic strengths with a global threshold θ on all the links). The threshold θ ranges from 1 to 10 % of the maximum strength over the dataset. The figure shows binarization for a representative dFC dimer. (**C**) The degree of temporal regularity in link activation/deactivation was assessed by quantifying the burstiness coefficient β, the mean activation time μ and the total activation time τ for every link and subject. The burstiness coefficient is bounded in the range -1 ≤ β ≤ 1 where it approaches to -1 if the link is tonic/periodic (blue lines), or it can approach to 0 if it has Poissonian (random-like) patterns of activation (red lines); β = +1 corresponds to links with bursty-like events of activation. (**D**) Distributions of β, μ and τ for the NC group, later used as reference. Upper and lower rows represent distributions over, respectively, intra zone and inter zone links (for an intermediate threshold, 0.0087 < θ < 0.0870). Left: Distribution of burstiness coefficients across different thresholds averaged over two subsets of intra- and inter-zone links. The β of intra-zone dimers approach to -1 and have more tonic/periodic patterns of activation (β = - 0.890 ± 0.027, median ± m.a.d), while the β inter-zone are closer to 0 and show more Poisson-like intermittency (β = -0.229 ± 0.020, median ± m.a.d). Middle: The mean duration μ which is bounded to the length of time-series for one subject (208 time-points), for the intra-zone links was longer than inter-zone links. Right: Analogously, the normalized total activation time (τ) of intra-zone links were longer than inter-zone links. (**E**) Mean values for the NC group were used as reference and percent relative variations were computed for the other SNC, aMCI and AD groups, combining relative values for different thresholds (see *Materials and Methods*). Upper and lower rows refer to intra- and inter-zone links. Left: Notice the large burstiness increase across groups for the inter-zone links (∼1.8% for aMCI and ∼9% for AD; green stars, p-value < 0.001; Mann-Whitney U-test) compared to a slight increase in the burstiness values of intra-zone links (∼0.5%). In contrast, SNCs showed a significant decrease of ∼ -6.5% relative to NC group in the inter-zone links. Comparisons between SNC, aMCI and AD for both intra- and inter-zone links were all significant (black stars). Middle: The mean activation durations of inter-zone links showed a relative negative decrease of roughly -1% for aMCI and AD subjects. Right: Total activation time τ was reduced to roughly -2% in aMCI and AD compared to NCs. Thus, temporal dynamics of dFC dimers are more tonic/periodic in SNCs than NCs and more intermittent in aMCI and AD subjects, particularly for inter-zone dimers.

In this approach, each link can assume up to four possible strength values, corresponding to its strengths in the FC^(λ)^ associated to each of the four states. Hence, any variability of dFC dimers reflects exclusively state-switching dynamics. Figure 3B shows the time-course for a representative fluctuating dFC dimer. The temporal organization of link fluctuations (periodic or bursty) can be highlighted by a binarization procedure, where a link is set to 1 if its instantaneous strength is above the threshold θ, or to 0 otherwise (see Materials and Methods). The result of this procedure is shown in Fig. 3C, for a few representative links and a specific choice of threshold. A link whose strength remains steadily above (below) threshold will result as constantly –or tonically– “active” (“inactive”). In contrast, a link whose fluctuating strength crosses the threshold through the different dFC-state frames will undergo several activation and inactivation events at specific threshold crossing times. Yet, there can be various types of intermittency, with different temporal statistical properties. The durations of different link activation and inactivation epochs could all be roughly similar, resulting in a more *periodic* type of intermittency (blue color link activation rasters in Fig. 3C). Alternatively, they could be more variable, stochastically alternating between shorter and longer activation epochs (red color rasters in Fig. 3C). The degree of temporal regularity in link activation and deactivation dynamics can be evaluated, link-by-link, by the quantification of a *burstiness coefficient* (β). We also define the mean duration of a link’s transient activation events as *mean activation* (μ) and the total fraction of time in which a link is active relative to imaging session duration, *total active time fraction* (τ). The burstiness coefficient is bounded in the range -1 ≤ β ≤ 1, with: β < 0, corresponding to near-tonic or periodic link activation dynamics; β = 0, corresponding to Poisson (random-like) link activation dynamics; and β > 0, corresponding to time-clustered (bursty) events of link activation. Mean activation times μ are bounded to the length of time-series. Total active time fraction is also bounded, 0 ≤ τ ≤ 1.

In this approach, three numbers β (burstiness coefficient), μ (mean activation) and τ (total active fraction) fully characterize the binarized dynamics of a link (for a given choice of the strength threshold θ). These metrics were evaluated for the two categories of dFC dimers: *intra-zone* (between two regions within either zone I or II) and *inter-zone* (between one region in zone I and one region in zone II). Our results show that these two categories have distinct distributions of β, μ and τ, first exemplified in NC subjects (Fig. 3D). Whereas Inter-zone dFC dimers are closer to a Poisson-like intermittency (β = -0.229 ± 0.020, median ± m.a.d), intra-zone dimers, present a tonic activation time-course (β = -0.890 ± 0.027, median ± m.a.d). In addition, inter-zone dimers are also less active (τ = 0.312 ± 0.099 for inter-zone vs. τ = 0.855 ± 0.027 for intra-zone dimers) and activate for shorter transient times (μ = 34.926 ± 4.439 for inter-zone vs. μ = 178.995 ± 7.378 for intra-zone dimers). These results suggest a smaller average strength of inter-zone time-averaged FC than for intra-zone FC. Using NC subjects as reference group, we measure indeed an average FC*(t)* strength = 0.083 ± 0.135 for inter-zone and of 0.564 ± 0.155 for intra-zone dimers (average ± s.d.). Similar differences were found for all groups (Table S1). The relative differences in β, μ and τ between intra- and inter-zone dimers are maintained over the entire range of possible thresholds θ (Fig. S1C for bustiness coefficient). Inter-zone dimers also displayed more burstiness, were more transient and less active than intra-zone dimers in all groups.

To achieve a robust and more precise comparison of β, μ and τ distributions between the cohorts (Fig. 3E), we computed percent changes of the three indicators in SNC, aMCI and AD groups relatively to normal controls. The advantage of relative comparisons is that they can be collated for different threshold values θ, resulting in a threshold-independent analysis. We found that, moving from NC to aMCI and AD subjects, many dFC dimer links tended to have larger burstiness values. In contrast, moving from NC to SNC subjects, dFC dimers tended to be more tonic. These trends of β were smaller yet significant for intra-zone FC dimers (Fig. 3E), compared to inter-zone dimers, reaching +1.869 ± 1.663 % for aMCI patients, +9.071 ± 3.001 % for AD patients and -6.404 ± 1.938 % for SNC subjects (Fig. 3E) that had larger values.

These results reinforce the notion of a significant reduction of inter-zone time-averaged FC along the clinical spectrum (cf. Table. S1). More importantly and beyond this reduction of average strength, our results point to a degradation of the temporal regularity of FC fluctuations. While the total active time fraction τ of inter-zone dFC dimers decreased by less than -2% from NC subjects to aMCI and AD patients (Fig. 3E; and even increased for intra-zone dimers), the burstiness of inter-zone links increased over 10%, showing a real alteration in the temporal statistics of link activation, well beyond the trivial decrease necessarily induced by the observed reduction of average strength.

We also observed a significant decrease of the mean activation time μ (Fig. 3E), for both intra-zone and inter-zone dFC dimers (−1.275 ± 0.227 % for aMCI and AD subjects compared to NCs). For SNC relative to NC, however inter-zone link burstiness decreased and their activation time increased (+0.613 ± 0.161 % for SNCs).

Goh and Barabasi (2008) also defined another metric related to burstiness, the memory coefficient. This coefficient λ (see Methods for exact definition) becomes significantly positive when autocorrelation exists in the duration of consecutive link activation events, i.e. when long-(short-) lasting activation events tend to be followed by activation events which also are long (short). Computing λ, we found a weak median autocorrelation in all four groups, for both intra- and inter-zone links. Values (see Supplementary Table S2) were small but still significant given the large number of activation events. Furthermore, memory was decreasing across the four groups from SNC to AD, providing yet another indication of increased disorder.

In summary, the temporal dynamics of dFC dimers between regions in different zones is altered along the SNC-AD spectrum from tonic and periodic in SNC to more intermittent in aMCI and AD subjects. Together with the finding of altered dwell times and transition dynamics between dFC states (Figs. 2C, D), our state-based dFC analyses based on the PBM approach suggest that changes towards AD involve a degradation of global integration and an increased disorderliness of dynamic functional interactions between zones.

### State-free dFC: entangled dFC flows in continuous time

The PBM approach to dFC analyses reduces the description of FC network reconfiguration to the tracking of discrete state switching events. Alternatively, sliding-window approaches evaluate the evolution of FC links as a continuous reconfiguration along time. As shown in Fig. 4A, all dFC dimers FC(*t*_1_) can be evaluated in a time-resolved manner restricting their estimation to BOLD signal time-series within a window centered at time *t*_1_. The window is then shifted at a slightly increased time *t*_1_ + δ*t,* providing an updated set of values FC(*t*_1_ + δ*t*). The result is a collection of smoothly varying continuous time-series FC(*t*) for each possible dFC dimer (Allen et al., 2014; Battaglia et al., 2020).

**Fig. 4.**
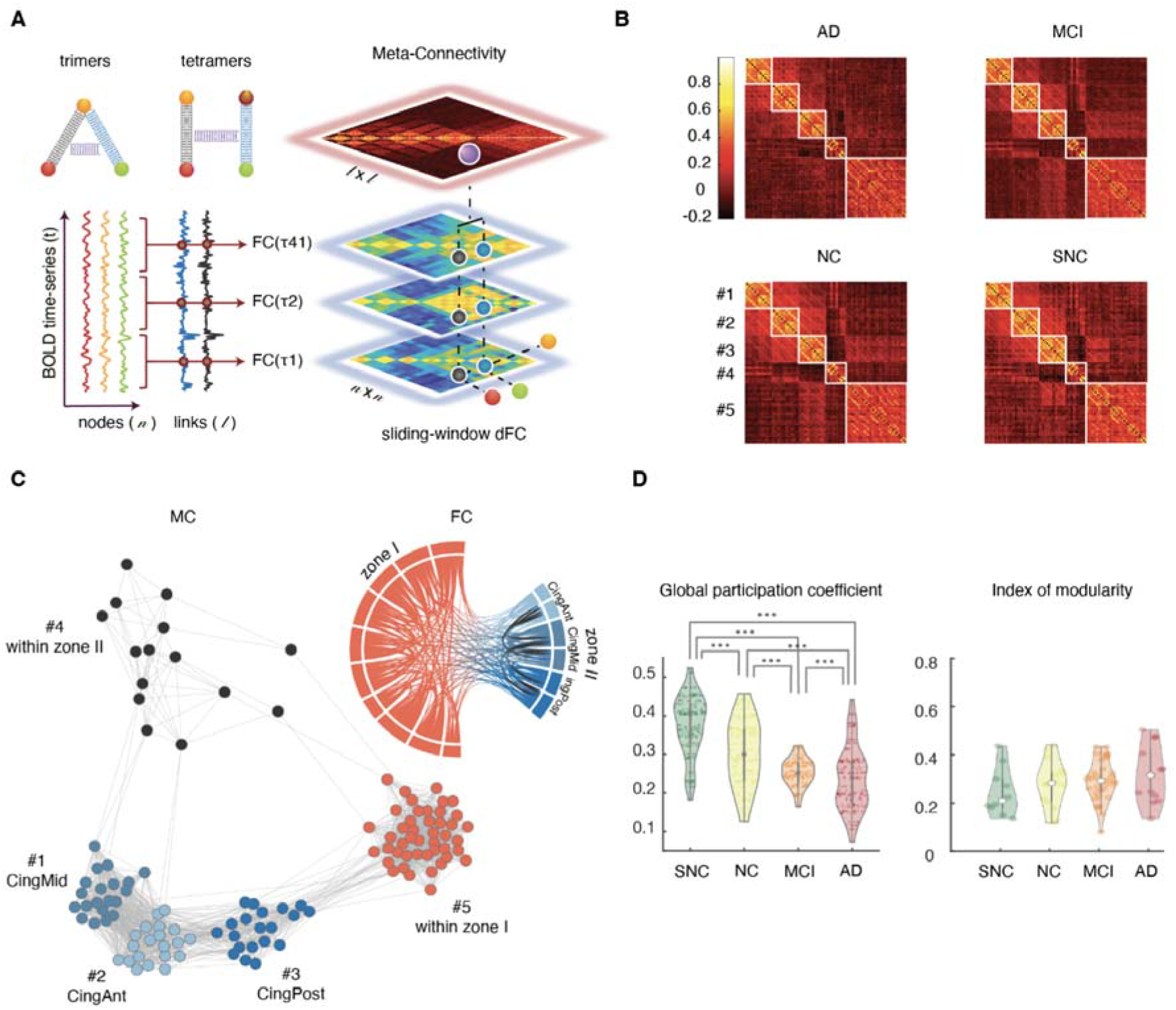
State-free dFC: Meta-Connectivity. **(A)** We slid a window of length = 5 TRs (10 s) with no overlap on the BOLD signals from the *n* considered regions. We then computed *n* x *n* FC matrices for each window using Pearson’s correlation between pair of regions. In this way each of the *l* possible pairwise links of FC becomes associated to a continuous time-series of varying FC strength. Correlations between these link time-series can be compiled in a *l* x *l* Meta-Connectivity (MC) matrix. We represent here trimer and tetramers with a spring between the involved dimers, as, in presence of meta-connectivity, pairwise links are not free to fluctuate independently. (**B**) Group average MC matrices for the four clinical groups. Louvain algorithm was applied on the MC matrices resulting in five modules. (**C**) A graph representation of the MC for the NC group, together with a chord-diagram of FC for the same group. Each node in the MC graph corresponds to a link in the FC graph. The different MC graph modules correspond thus to different types of links: MC modules #1, #2 and #3 include inter-zone links incident, respectively, to medial, anterior and posterior cingulate cortices (edges within these modules are thus inter-zone trimers rooted in Zone II); MC module #4 and #5 include links, respectively, within zones II and I. (**D**) Modules are also connected between them. The relative amount of inter-module meta-links is captured by the global participation coefficient (averaged over the five modules) which showed a significant decrease across the clinical groups (Mann-Whitney U-test, p < 0.001).

As in the case of node activity time-series, it is possible to study covariance between the temporal evolutions of different dimers. The case in which their fluctuations are not independent –or, in other words, that the dimers are “entangled”– will be signaled by significantly positive or negative correlations between dimers. These correlations can be represented graphically by trimer and tetramer diagrams in which the two entangled dimers are linked by a spring (Fig. 4A, top left; we will omit in the following to draw this spring, for the sake of a clearer visualization). The stronger the correlation between the fluctuations of different dFC dimers in a trimer or tetramer, the stronger will be their “entanglement” (i.e., metaphorically, the stiffness of the spring).

These strengths of entanglement between FC dimers can be compiled into a *meta-connectivity* matrix (MC; Fig. 4A). The notion of MC (Lombardo et al., 2020) is strongly related to the edge-centric FC discussed by Faskowitz et al. (2020). The key difference is that MC is obtained by using a short smoothing window in the estimation of the stream of FC(*t*) matrices, while edge-centric connectivity captures coincidences between instantaneous fluctuations. The denoising brought by the smoothing window allows an easier extraction of the modular structure of MC, with respect to edge-centric FC (cf. Lombardo et al., 2020), but the two concepts are otherwise equivalent. The choice of window size (here 5 TRs, *Materials and Methods*) was motivated by the fact that the state-based PBM method suggested that ∼90% of epochs within a coherent state lasted less than 5 TRs (Fig. S2A), indicating a fast intrinsic timescale of link fluctuation. Furthermore, we can observe *post-hoc* that the use of larger (or smaller) windows would not improve the capability to separate our groups based on MC values (Fig. S2B).

Group-averaged MC matrices are shown in Fig. 4B for the four groups. Their modular structure is evident at simple visual inspection. A module in the MC matrix –also called dFC module or meta-module (Lombardo et al. (2020))– corresponds to a set of co-fluctuating dynamic FC links, i.e. to FC subnetworks whose overall strength waxes and wanes transiently along the resting state in an internally synchronous manner. The existence of non-uniform MC matrices indicates that the flow of dFC reconfiguration is not mere noise but rather, it is organized by specific arrangements of “springs between the links”. In other words, fluctuations of FC dimers are entangled in complex patterns reflecting higher-order correlations (non-vanishing trimers and tetramers) between the coordinated activation of multiple regions.

### dFC flow in patients is less globally entangled

MC matrices can also be represented as graphs, in which MC-nodes correspond to different FC-links and MC-links appear due to the entanglement between the FC-links. An example graph embedding is shown in Fig. 4C for the MC matrix of the NC group. Graph vertices are color-coded depending on the type of associated FC link (i.e. start and end zones of the links, cf. FC diagram with matching colors at the top right of Fig. 4C). Notably, the different dFC modules, visible as blocks in the MC matrices of Fig. 4B and as uniform-color node communities in the graph of Fig. 4C, are composed of FC dimers with internally homogeneous start and ending zones.

A standard graph-theoretical notion useful when commenting about dimer arrangements into trimers and tetramers is the one of *incidence:* a link is incident to a node (or a node incident to a link), if the link is attached to the node (the notion of incidence complements the more familiar one of *adjacency,* where two nodes are said to be adjacent if connected by a link). Equipped with this terminology, we call *root* the common region incident to both the dimers within a trimer, while the other two regions form the *leaves* of the trimer. We can then describe the first three dFC modules (#1, #2 and #3) of the MC matrix as including mutually entangled FC dimers originating in either one of the Zone II cingulate regions and terminating in Zone I. The entanglement of FC dimers gives thus rise to strong inter-zone trimers with “roots” in Zone II and “leaves” reaching out to Zone I regions. The two other dFC modules #4 and #5 include dimers within Zone I and Zone II, respectively, forming strong within-zone trimers or tetramers. Entanglement is thus particularly strong between dimers within a same zone and between inter-zone dimers incident on a common root region (in Zone II).

Although the MC graph is highly modular, it is not split into disconnected components and some entanglement exists also between dimers located in different dFC modules. Inter-module connections in the MC graph can arise e.g. due to the existence of trimers with a root in zone I (entangling dimers across dFC modules #1, #2 and #3) or inter-zone tetramers (entangling dimers across dFC modules #4 and #5). In other words, MC reveals some degree of global, widespread entanglement between FC dimers, beyond modular entanglement. The strength of such global entanglement is quantified by the so-called average *participation coefficient* of the MC matrix, a graph-theoretical quantity measuring inter-module coupling (Guimerà & Amaral, 2005; see *Materials and Methods*).

The distribution of MC participation coefficients for each group are shown in Fig. 4D. We found that the participation coefficients decreased significantly (Fig. 4D, left; Mann-Whitney U-test, p <0.001) from SNC to AD, while overall modularity did not vary significantly (Fig. 4D, right). These results suggest that, in patients, coordination structure between fluctuations of FC dimers is impoverished: global entanglement is disrupted, making dimer fluctuations in different modules more random and mutually independent.

### Interlude: trimers and tetramers are genuine or “dimers are not enough”!

Before entering a more detailed and regional specific account of changes to dFC organization observed at the regional level along the SNC-to-AD spectrum, it is important to stress that trimer and tetramer analyses are *not redundant* with the dimer-based analyses. Indeed, some studies have suggested that correlation between edges (captured by higher-order trimer and tetramer in a MC matrix) could just be an automatic byproduct of existing lower-order dimer interactions (Novelli and Razi, 2022). This can be easily understood through some examples. Let consider for instance two strong dimers FC*_ri_* and FC*_rj_* sharing a common root region *r*. If a third strong dimer FC*_ij_* also exists –closing the triangle of edges *(ri), (rj), (ij)*, then it is not surprising that a strong trimer MC*_ri, rj_* is also detected: indeed, the fluctuations of the two leaf regions *i* and *j* are coordinated through a transverse dimer interaction, i.e. the strength of the trimer would be the byproduct of a triangular motif of dimers and would thus be a redundant consequence of them. Analogously, we may consider the case of a square motif of dimers FC*_ij_*, FC*_jk_*, FC*_kl_* and FC*_li_* which could also give rise to strong tetramers because of the presence of one or more pairs of strong dimers. In other words, the detection of strong trimer and tetramer entries within the MC (or other forms of edge-centric FC) is not a sufficient condition for the existence of genuine high-order interactions (Battiston et al., 2020) that cannot be explained as stemming from motif arrangements of lower-order pairwise interactions. On the contrary, the existence of genuinely high-order interactions could be established by detecting trimer or tetramer couplings between the dimers in a motif, stronger than the dimers themselves involved in the motif. The question that then arises is, what is the structure of MC that we observe in our data?

To investigate the genuine or spurious nature of trimer and tetramer interactions, we systematically studied the inter-relations between MC and FC entries. First, we define the *dimer strength* FC*_r_* = Σ*_i_* FC*_ri_* of a region *r* as the sum of the strengths of all the dimers incident to it. Analogously, we introduced the (root-pinned) *trimer strength* MC*_r_* = Σ*_ij_* MC*_ri, rj_* of a region *r* as the sum of the strengths of all the trimers of which *r* is the root. Conceptually, whereas FC*_r_* measures the average coordinating influence that the region *r* exerts on its adjacent nodes, MC*_r_* can be understood as quantifying the coordinating influence that *r* exerts on its incident links. As shown by Fig. 5A, the correlations between dimer and trimer strengths of a region are weak and not significant, both at the global (black lines) and within each group (bundles of colored lines) levels, and for both within-zone and inter-zone trimers and dimers strengths. Of note, the average strength of between-zone trimers and dimers strengths had a larger variance across groups, hence the positively slanted shape of the global point cloud when confounding all groups, despite negative trends within each group. Although weak, within-subject correlations between FC*_r_* and MC*_r_* were negative, suggesting that some regions can be “meta-hubs” (Lombardo et al., 2020) but not “hubs”, i.e. they can be the center of an entangled star subgraph of incident dimers, even if these dimers are individually weak and unable to systematically synchronize the fluctuations of adjacent nodes. Such meta-hubs could not have been identified through ordinary pairwise FC analyses only and manifest thus the existence of a real high-order multi-regional coordination.

**Fig. 5.**
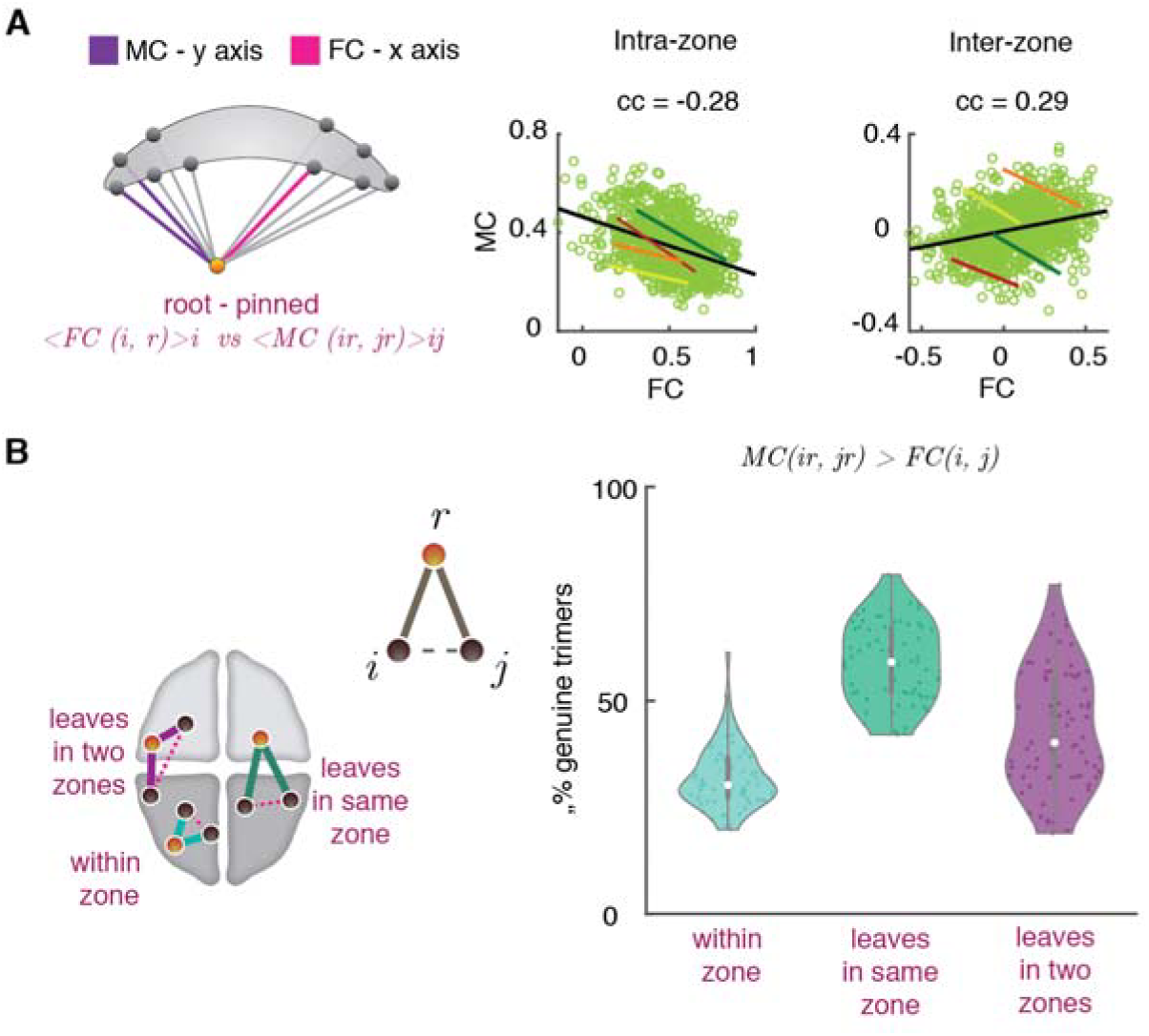
State-free dFC: Inter-relations between dFC trimers and FC dimers. We studied whether regions with a large FC strength (“FC hubs”, i.e. they are the center of a star of links strong on average) also have a large trimer strength (MC “meta-hubs”, i.e. they are the center of a star of links whose fluctuations are temporally correlated). (**A**) To do so we computed the correlation between dimer and FC strengths, for both within and between zones trimers and dimers. As shown by the scatter plots, these correlations were low, both at the global (light green cloud) and at the single clinical group (colored solid lines; green: SNC, yellow: NC, orange: aMCI, red: AD) levels. Within each group, they were furthermore moderately negative. Therefore, FC hubness and MC meta-hubness tend to be slightly anti-correlated. (**B**) Trimers were divided into three groups dependent on the location of their *roots* and *leaves*. We considered *genuine* a trimer such that the MC between the two dimers composing the trimer is stronger than the FC between the trimer leaves. The violin plots at the right show fractions of genuine trimers (for all trimers and subjects) as a function of the trimer type. For all types, there were substantial fractions of genuine trimers (i.e. higher-order interactions not fully explained by the underlying dimer interactions arrangement). See Figure S3 for analogous analyses on tetramers.

We then moved to consider how many trimers cannot be considered as a manifestation of underlying triangular motifs of dimers. We defined a trimer rooted in a region *r* to be *genuine* if MC*_ri,rj_* > FC*_ij_*, i.e. if the observed trimer strength cannot be fully explained by a strong synchronization between the leaves. We then measured the observed fractions of genuine trimers. As shown by Fig. 5B, substantial fractions of genuine trimers could be found for all trimer types: genuine fractions amounted to 32 ± 7 % for *within zone trimers* (root and both leaves in a same zone) and increased to 43 ± 13 % for *inter-zone trimers* with *leaves in two different zones*, or 58 ± 9 % for *inter-zone trimers* with the *root in a different zone than the leaves*. Especially for inter-zone trimers, many trimers could not be trivially explained by the existence of triangles of dimers.

Considering tetramers, we found larger redundancy with dimers. We defined the *tetramer strength* MC*_ij_* = Σ*_kl_* MC*_ij, kl_* of a link (*ij)* as its total entanglement with other links. Figure S3A shows that a significant positive correlation existed between the dimer strength FC*_ij_* of a link *(ij)* and its tetramer strength. That is, the stronger links were also the most entangled. Interestingly, several tetramers could still be considered genuine. We defined a tetramer genuine when MC*_ij,kl_* > FC*_ij_*, i.e. when the two composing dimers were strongly correlated, despite (at least one of) the dimers being individually weak. Under this definition, Figure S3B shows that up to 55 ± 10 % of tetramers composed of interzone dimers were genuine.

We conclude that in general, the information conveyed by trimer and tetramer analyses is not completely redundant with the one conveyed by dimers, as many trimer and tetramer metrics cannot be explained solely in terms of dimers and thus express actual higher-order correlations.

### dFC trimers and tetramers are more impacted in aMCI and AD than FC dimers

After defining various metrics to quantify the involvement of specific regions and links into pairwise and higher-order interactions, as previously described, we then studied how dimer, trimer and tetramer strengths varied across the four cohorts in our study.

First, we found that for both dimer and trimer interactions, the stronger effects were found considering inter-zone interactions. Figure 6A reports group differences for inter-zone dimers and Figure 6B for inter-zone trimers (mixed-zone or same-zone leaves are not treated separately). Results for within-zone dimers and trimers are shown in Figures S4A and S4B, respectively. In contrast to within-zone interactions, group-level comparisons for within-zone dimer and trimer interactions were not significant.

**Fig. 6.**
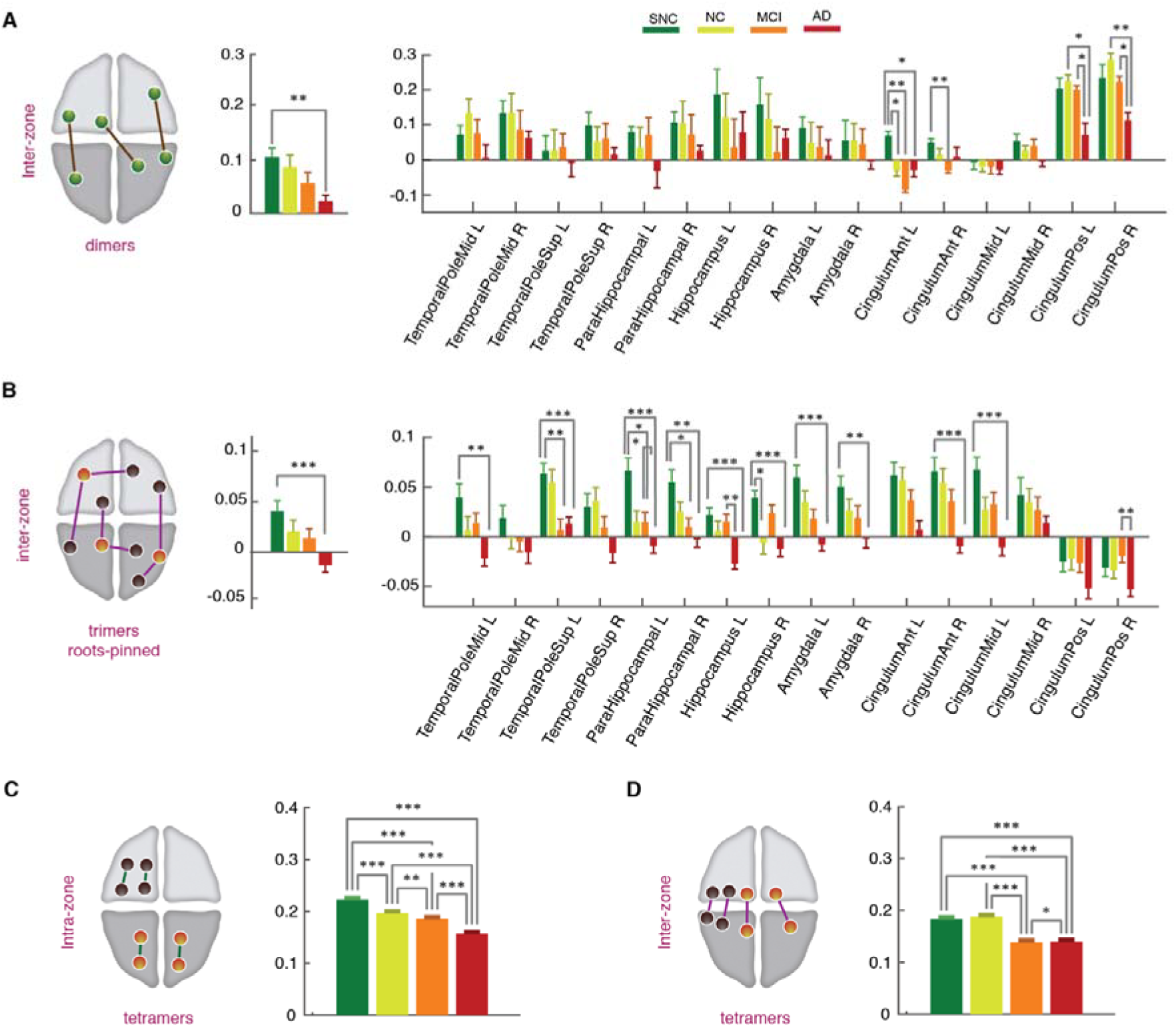
State-free dFC: strengths of inter-zone FC dimers, trimers and tetramers across clinical groups. (**A**) Average strength of inter-zone FC dimers decreased from SNC-to-AD both globally (left) and locally at the level of individual regions (right). At the global level, significant differences were found between the SNC and AD groups (p-value = 0.005, Mann-Whitney U-test, Bonferroni correction). Locally the decrease was significant in anterior and posterior cingulate gyrus, bilaterally (Mann-Whitney U-test, Bonferroni correction). (**B**) Inter-zone trimer strengths, similarly to FC dimers, showed a reduction trend across the groups, both globally (left) and locally (right). At the regional-level the reductions in dFC trimers were widespread among regions, including early-affected regions without noticeable FC strength variations across clinical groups, with an interesting tendency toward negative trimer strengths in the AD group, associated to developing “frustration” of higher-order interactions in a statistical mechanics sense (and, correspondingly, increased dynamical disorder and conflict; see *Discussion*). Finally, **(C-D)** tetramers strength showed a significant drop from SNC to AD groups in both brain-wide averaged intra-zone (**C**) and inter-zone (**D**) subsets. See Figure S4 for intra-zone dimer and trimer strengths, not showing significant variations across groups.

In general, when averaging over all brain regions (Figs. 6A and 6B, left), general averages of dimer and trimer strengths progressively decreased from SNC, to NC, aMCI and AD groups. This decrease, notably, was significant when comparing the two extreme SNC and AD groups. The effect was particularly strong for inter-zone trimer strengths (p = 0.005, Mann-Whitney U-test, Bonferroni correction, for trimers), whose average value for the AD group not only decreased but changed its sign as it became negative. In contrast, within-zone trimer strengths remained strongly positive (Fig. S4B). This means that, in the AD group, several regions are involved in a mixture of negative and positive trimer interactions. Positive interactions tend to synchronize the fluctuations of FC links, unlike negative interactions that tend to push them in an anti-phase interaction. Furthermore, the mixture of positive and negative couplings results in a dynamic conflict scenario, known in the statistical mechanics as *“frustration”* (Vannimenus and Toulouse, 1977) and has been associated to disordered organization and a slowed-down relaxation to equilibrium (Mezard et al., 1988). The emergence of frustrated inter-zone trimer interactions is a strong qualitative discriminative marker of the AD group (see *Discussion* for possible interpretations of this finding).

The decrease of inter-zone trimer-strengths and their switch to negativity in the AD group is confirmed also when focusing on individual brain regions, rather than the average (Figure 6B, right). Remarkably, strong decrease in trimer strengths were observed in regional subdivisions of the Temporal Pole and of the Parahippocampal gyrus, along the Hippocampus proper and Amygdala. Some of these regions (Entorhinal cortex in the Parahimpocampal gyrus and the Hippocampus), are among the first to be affected by neurofibrillary accumulation in AD pathology (Braak stages 1 and 2). In these same regions, we found a similar trend at the level of dimer strengths even when differences were not significant (Figure 6A, right). Of interest, the stronger effects at the level of dimer strengths were found in the Cingulate gyrus which are affected by early beta amyloid depositions and later on with neurofibrillary accumulation. Interestingly, the regions exhibiting the strongest effects at the level of trimers were not the ones with the strongest effects at the level of dimers (and vice versa; Fig. 6A right vs Fig. 6B right). The two analyses reveal thus complementary aspects of how pathology affects the spatiotemporal organization of functional interactions.

Lastly, we assessed differences on tetramer strengths across groups. In Figure 6 we show the average tetramer strengths for intra-zone (Fig. 6C) and inter-zone (Fig. 6D) tetramers. In both cases, we observed a significant reduction of tetramer interactions from the SNC, to the NC, MCI and AD groups. In the case of inter-zone tetramers, the drop in strength was large in the MCI group, with levels close to those in the AD group.

In summary, AD was associated with extensive reductions of not only dimer strengths, but more importantly, trimer and tetramer strengths. Furthermore, inter-group differences were salient when considering higher-order trimer and tetramer compared to dimer interactions.

## Discussion

We have shown a large variety of changes associated with dFC across the cognitive spectrum from cognitively over-performing SNC subjects to AD. The rich set of complementary analysis approaches we deployed consistently converge toward a common message: AD is associated with a disordering of the rich spatiotemporal fluctuations that characterize healthy dFC.

It is worth noting that while BOLD activity misses many fast neuronal processes due to its slow sampling rate, what Functional Connectivity dynamics track are not neural level processes but variations of *global brain state* that can occur on much slower time-scales. So dFC with a long TR accounts for variations of the way in which the repertoire of internal states is sampled, more than for variations of neural signals themselves. As a side note, these slow fluctuations are also what mean-field connectome-based whole-brain models are fit to reproduce via the stochastic sampling of their emergent repertoire of dynamic modes (Hansen et al., 2015, Fousek et al., 2022).

Our results showed that a pertinent description of dFC organization and its changes across groups can be formulated in terms of two anatomical zones segregating ventral from dorsal areas (Fig. 6D). We found that the system spends less time in states with fluid Zone I dynamics and high global integration, visiting them more transiently, while it gets stuck on the contrary in less integrated states exhibiting Zone I hypersynchronisation (Fig. 2). At the dimer level, pairwise interactions between regions in different zones get more irregularly bursty (Fig. 3). At the level of higher order trimers and tetramers, meta-connectivity analysis revealed a loss of coordination between the fluctuations of different sets of links, as quantified by dropping participation coefficients (Fig. 4D). Trimer interactions between Zone I and Zone II, as well as tetramers, were weakened more distinctively than the inter-zone dimer interactions. Remarkably, regions in our limbic subnetwork for which conventional dimer analyses were not different between groups, showed a remarkably reduced involvement in trimer interactions between zones (Fig. 6). Overall, these findings point together toward a “loss of structure” in dFC in parallel to the cognitive gradient across groups. This is in agreement with previous studies that showed a reduction of the complexity of spontaneous fluctuations of coordinated activity (Tait et al., 2020).

Nevertheless, even though being quite encouraging, a conclusive validation of our findings would require using larger cohorts, which preferably contains information on cortical thinning and PET scans of tau and Aβ depositions, to test whether their distributions correlate with the local network dynamics alterations we observe (thus establishing them as potential physiopathological causes of these changes) or not (advocating for alternative explanations, see later discussion). Similarly, our choice of regions and parcellations was arbitrary, generally based on the successful use of the same parcellation in previous modelling-based analyses of the same cohort (Zimmermann et al., 2018b). A better resolution fMRI from further cohorts would allow validating our results with finer and more extended parcelations, especially for the subcortical regions (Tian et al., 2020) that constitute the core of the limbic network on which we have focused.

Interestingly, our qualitative description emerges from radically different approaches to dFC parameterization: a state-based approach (the PBM method by Thompson and Fransson, (2016)); and a state-less approach (the random walk descriptions of dFC by Battaglia et al. (2020) and Lombardo et al. (2020)). The PBM method is firmly rooted in the developing field of *temporal network theory* (Holme and Saramäki, 2012). Temporal networks allow describing inter-regional communication as it unfolds in time, similarly to a call-center, where operators can handle a multitude of brief first-contact calls at certain moments and dedicate extensive time to select customers at other times (Kovanen et al., 2013). Or to a primary school, where students interact in small groups during lectures and play in mixed larger groups in the playground during school-breaks (Gemmetto et al., 2014). Eventually, even fluctuations between segregated or integrated states in brain systems at different scales (Shine et al., 2016; Pedreschi et al., 2020) give rise to network dynamics not dissimilar to these social systems. Note that our use of terms such as “burstiness” or “activation” (cf. Fig. 3D and E) is also mediated from the jargon of temporal networks theory and should not be mistaken with the usual meaning of these terms in neuroscience, as they refer to FC link dynamics rather than to neuronal firing rates (exactly as we use the adjective “temporal” in the sense of “time-dependent” and not in association with “temporal lobe”).

The dFC random walk approach (Arbabyazd et al., 2020; Battaglia et al., 2020; Lombardo et al., 2020; Petkoski et al., 2023) models rs dFC as a temporal network as well, but focuses on the variation from one network frame to the next, more than on the geometry of individual network frames. dFC is seen as a flow in network space and the non-randomness of network reconfiguration was investigated via a time-to-time correlation approach known as Meta-Connectivity (Lombardo et al., 2020). In a dFC context in which the mode of coordination between regions is not frozen in time but changes smoothly, meta-connectivity reveals how the fluctuations of one or more regions modulate the degree of coordination between the fluctuations of other regions. In other words, meta-connectivity is an indicator of “many-body coordination”. Indeed, the terminology of dFC “dimers, trimers, tetramers” is reminiscent of perturbative diagrammatic expansions in Statistical Physics, such as the virial expansion (Landau and Lifshitz, 1980), in which clusters of increasingly large size account for progressively more elaborate and nonlinear patterns of many-body interactions. MC can thus be considered yet another form of high-order functional connectivity, adding up to a list of other approaches to track higher-order coupling (Torres et al., 2021; Santoro et al., 2023) as hypergraph or homological methods (Battiston et al., 2020; Petri and Barrat, 2018; Sizemore et al., 2018), which have already identified synergistic aspects of human brain functioning (Luppi et al., 2022; Varley et al., 2023).

Unfortunately, both of the dFC methods implemented in this study provide results depending on specific parameter choices. For instance, concerning the state-less random walk approach, the selection of a window-size remains ultimately arbitrary. The window-size selected was short in contrast to other studies. However, our statistical analyses suggest that this window size results in similar discriminatory power as longer windows (Fig. S2A). Furthermore, it is necessary to use short windows because the PBM method suggests that dwell-times in consistent FC state epochs are often short and thus dFC is intrinsically fast (Fig. S2B). The need to track the covariance of fast FC fluctuations has inspired additional approaches analogous to MC, as edge-centric Functional Connectivity (eFC; Faskowitz et al., 2020). In this approach, covariance is estimated between individual events of instantaneous co-fluctuation, without arbitrary windowing. However, we showed in Lombardo et al. (2020) that, despite the significant relation between MC and eFC, the use of a sliding-window in the MC approach produces a smoothing effect that partially denoises the graph structure of inter-link meta-connections, allowing a cleaner determination of modules and “meta-hub” nodes with large trimer strengths.

An additional aspect of the state-based PBM approach, is that it involves partially arbitrary steps as the choice of a number of states. The retrieved FC states depend on the extracting algorithm that depends on the brain parcellation and choice of regions of interest utilized. We found four states and increased dwell-times in states with hyper-connectivity within Zone I. This finding of increased probability in AD of visiting hyper-connected states is in agreement with some state-based dFC studies (Gu et al., 2020), but in contrast with others (Fu et al., 2019; Schumacher et al., 2019), which instead find higher dwell-times in disconnected states. Such discrepancies may arise because in the PBM method clustering of states is performed on activation patterns rather than on time-resolved functional networks. Our procedure has the advantage of showing that network dynamics is partially dissociated from node dynamics, with the possibility of hyper-connected FC modules arising both in presence of higher or lower activity of the nodes composing this module (Fig. 2A). It may reduce the chance, however, of detecting extreme events along dFC or transient atypical network configurations that would be naturally assigned to separate clusters when directly clustering networks. Finally, the mentioned studies used reference parcellations with a larger number of regions or focusing on more distributed network components, while here we particularly emphasize selected regions of interest, such as temporal and paralimbic cortices, known to develop epileptiform activity (Bakker et al., 2012; Cretin et al., 2016; Vossel et al., 2013). Thus, within the probed sub-system of interest, hypersynchrony may become particularly prominent and over-expressed (hence, the enhanced dwell-time in hyper-connected FC states), a fact that has direct pathophysiological relevance.

Despite the arbitrary steps involved, both approaches independently provide sets of results with a high mutual consistency, making unlikely that our analyses reflect exclusively methods artefacts. Both methods confirm indeed that a dFC description in terms of two zones is pertinent, as the distinction between Zones I and II organizes the modular structure of both FC states in the state-based PBM approach (Fig. 2A) and of the MC matrices in the state-free dFC random walk approach (Fig. 5B and C). Furthermore, both methods confirm that the increased severity of cognitive decline across the four groups correlates with a reduced inter-zone coordination: more time spent in states with weaker integration (Figs. 2B-C) and reduced inter-zone trimer strengths (Fig. 6B). Such semantic agreement is remarkable especially given the limitations of our approaches. Meta-connectivity analyses could be improved by seeking, beyond plain module detection, for a hierarchical community structure, that is often present in large networks (Jeub et al., 2018; Peixoto, 2014). State-based analyses could profit of better clustering approaches, as used by Rasero et al. (2018). However, while acknowledging these limitations, we found our four states and MC communities to be already highly interpretable, in term of the anatomical nature of the entangled links.

Particularly interesting is the fact that the weakening of inter-zone trimer interactions across the four groups decreases to such extent that some of these trimer switch from a positive to a negative value. As previously mentioned, the coexistence of negative and positive couplings in a graph or a hypergraph of interacting units is referred to in statistical physics as “frustration” (Toulouse, 1986), since it is associated with the emergence of conflicts preventing smooth relaxation to an equilibrium. To put these results in context, let us imagine that a dynamic FC link (a dimer FC*_ij_*) is positively coupled to a second dimer FC*_kl_* and negatively coupled to a third dimer FC*_mn_*, and that the second and the third dimer simultaneously increase in strength (i.e. FC*_kl_* and FC*_mn_* get larger). Then the dynamics of FC*_ij_* will “freeze” under the contrasting influence of the positive bias applied by FC*_kl_* (pushing it to assumer stronger values), and the negative bias applied by FC*_mn_* (pushing it to assume smaller values). Thus, the change of positive to a negative inter-zone influence –as the one signaled by the negative inter-zone trimer strengths of many limbic region within Zone I– gives rise to conflicts between the flows of Zone I and Zone II regions in AD patients, in contrast to control subjects where the fluctuations of the same regions are naturally synchronized.

In particular in the context of cognition, Zone II regions such as the posterior Cingulate Cortex (pCC) have been postulated to play a regulatory role on the level of brain meta-stability, balancing “free-wheeling” internal cognition and focused outward attention (Leech et al., 2012; Leech and Sharp, 2014). In control groups, pCC has strong positive dimer coupling and moderately negative trimer coupling with regions in Zone I (Fig. 6). This could allow the pCC to quickly coordinate with individual Zone I regions (and share information with them via direct positive FC dimers), while simultaneously “lowering the volume” of intra-zone I communication (via pCC-rooted negative trimers with Zone I leaves). In AD subjects, this subtle equilibrium is lost, resulting potentially in perturbed integration of information within and between Zone I regions. Remarkably, pCC is also a key hub of the Default Mode Network (Raichle et al., 2001), a system whose dFC had already been suggested as a biomarker in the conversion to AD (Jones et al., 2012; Puttaert et al., 2020).

Interestingly, our analyses on trimer strengths could detect inter-group differences within Zone I regions, for which the dimer analyses did not found significant differences. A possible explanation for the better sensitivity of trimer-based analyses could trivially be due to a larger sample-size, as there were more possible trimers than dimers, resulting in similar average strengths but with a lower variance. However, another possibility could be that higher-order interactions are readily affected by the pathology process earlier or at a higher degree than pairwise interactions. This fact is difficult to assess from our dataset, which is not longitudinal. Yet, this possibility is supported by our results showing that higher-order trimers and tetramers terms convey in many cases genuinely new information, not redundant with dimer analyses. Indeed, even if we agree with other reports (Novelli and Razi, 2022) that dimer terms can sometimes explain trimer and tetramer term, we found in addition important trimer entanglement among otherwise individually weak dimers (Fig. 5A) that lacked strong pairwise interactions between their dangling leaves (Fig. 5B). Such genuine trimers cannot be explained by dimer motifs and describe thus a qualitatively different phenomenology, invisible to conventional FC analyses. Similar considerations apply to tetramers (Fig. S3), which although generally weaker in strength than dimers and trimers, form an additional and pervasive background “medium” which also actively steer coordinated FC dimer fluctuations, with an overall influence degraded by the pathological process (Fig. 5C and D). In the future, for an even better appreciation of pathology effects on higher-order interactions, one may use methods that facilitates the generalization to arbitrarily high orders, even higher than the third or the fourth one, such as maximum entropy fitting (Ezaki et al., 2018; Savin and Tkačik, 2017) or other information-theory approaches (Rosas et al., 2019).

Another question is what the mechanistic origin could be of the observed spatio-temporal complexity of dFC (and of its alterations). Previous studies have shown that structured dFC may emerge as an effect of global brain network dynamics to be tuned at a slightly subcritical working point (Arbabyazd et al., 2020; Glomb et al., 2017; Hansen et al., 2015), or as a consequence of cascades of neuronal activations (Rabuffo et al., 2021) that occur due to the flow on the manifold created by the symmetry breaking of the connectome (Fousek et al., 2022). However, these studies did not use very precise criteria when referring to their capacity to render dFC. In the future, the statistical descriptors of dFC alterations that we introduce here, such as regional spectra of trimer and tetramer strengths, may be used as more detailed fitting targets for the tuning of mean-field models aiming at explaining the circuit mechanisms for the emergence of higher-order interactions. Such models, once fitted, may also allow reverse-engineering the physiological changes that are responsible for the degradation of spatiotemporal dFC complexity along the SNC-to-AD spectrum.

It is likely that the dFC alterations we observe between groups are caused at least in part by underlying biological causes of AD, as the aggregation of misfolded proteins that cause cell death and atrophy (Soto & Pritzkow, 2018). However, not all the symptoms can be explained by these mechanisms. Among them, the existence of symptom severity fluctuating across hours in a way not accountable for sudden variations of amyloid load (Palop et al., 2006) or, yet, the phenomenon of cognitive reserve where subjects with virtually identical or even higher amount of amyloid load than others can maintain a very efficient cognition, (cf. Snowdon (2005) for the famous “Nun Study” or Rentz et al., (2010) for a review of other studies with similar conclusions). These findings suggest that neurodegeneration may coexist with compensations of unspecified nature that allow “cognitive software” to operate properly despite “hardware damage” (see e.g. Petkoski et al. (2023) for examples of dynamic compensation in healthy aging, or Courtiol et al. (2020) for a similar phenomenon in epilepsy). Here, we propose the hypothesis that preserved dFC complexity may act as a possible form of cognitive reserve. We stress once again that, to check the soundness of this hypothesis, future analysis should rely on richer datasets that contain PET scans of tau and Aβ depositions, and possibly even a mechanistic model (Stefanovski et al., 2019; 2021) for their impact to the neuronal activity.

Ultimately, the degradation of dFC organizational complexity that we here described may not only correlate with cognitive decline but also, eventually, contribute to cause it. Indeed, a dFC with a complex organization could be the hallmark of brain dynamics implementing “healthy” cognitive processing. Computation can emerge from collective dynamics as long as this dynamics is sufficiently complex, i.e. neither too ordered nor too random (Crutchfield, 2012; Crutchfield and Mitchell, 1995). More fundamentally, the existence of alternative information processing states –transient FC networks?– and of non-random transitions between these states –structured and complex dFC switching?– are two necessary conditions for whatever information processing system to perform computation (Turing, 1937). A speculative hypothesis is thus that the complexity of neural dynamics –and, more specifically the complexity of ongoing dFC which is a measurable shadow of hidden neural processes– is an instrumental resource for cognitive information processing. Cognitive deficits in pathology could arise just in virtue of this resource becoming scarcer, because of less structured and more random dynamics. This phenomenon has been speculatively observed in hippocampal neuronal assembly dynamics in epilepsy (Clawson et al, 2021). In this line of thinking, preserved dFC complexity would act as a “dynamic reserve” allowing the implementation of elaborate neural computations (or “software patches”) to compensate for progressing neurodegeneration. Analogously, enhanced dynamic complexity could be the substrate for the superior cognitive performance achieved by subjects in the SNC group with respect to NC subjects. A more direct exploration of the link between dFC complexity and cognitive processing in the healthy and pathological brain will be needed to inquire into this suggestive hypothesis.

## Materials and methods

### Participants

The study included 73 subjects between 70 and 90 years of age from the fourth wave of the Sydney Memory and Ageing study (Sachdev et al., 2010; Tsang et al., 2013). The use of the database was approved by the Human Research Ethics Committee of the University Texas at Dallas. For detailed descriptive summaries on neuropsychological assessments for AD and amnesic aMCI, we refer the reader to Zimmermann et al. (2018).

A specificity of our approach is the stratification of healthy controls with an additional “super normal” category putting our focus not only on mechanisms of disease but also on mechanisms of “health” based on cognitive performance. Results from twelve neuropsychological tests were combined in the following cognitive domains: attention/processing speed, memory, language, visuospatial ability, and executive function. In brief (Mapstone et al., 2017) we classified cognitive membership for each subject based on the composite Z-scores as supernormal controls (SNC) or normal controls (NC). For this, the supernormal (SNC) group was defined as *Z_mem_* > 1.35 SD (∼90th percentile) and *Z_cog_* > 0.7 SD. The normal control participants are conservatively defined with *Z_mem_* ± 0.7 SD (∼15th %ile–85th %ile) of the cohort median. The classification of subjects as AD and aMCI described in Zimmermann et al was done by consensus included the following: The amnesic MCI group was described by a cognitive decline at least in the memory domain (*Z_mem_* and/or *Z_cog_* < 1.5 SD below normative values), paired to subjective complaint of cognitive deficit and without deficits in activities of daily living (ADL). The AD group in presence of a diagnosis of Alzheimer’s Disease according to DSM-IV criteria (American Psychiatric Association, 2000) assessed by a clinical expert panel that included significant cognitive decline in several cognitive domains in addition to significant decrease in ADLs (American Psychiatric Association, 2000; J. Zimmermann et al., 2018).

### fMRI acquisition and preprocessing

Details about resting state functional MRI acquisition and preprocessing can be found in Zimmermann et al. (2018). We briefly mention, as relevant here that during the fMRI acquisition, participants were instructed to lie quietly in the scanner with their eyes closed. The TR used for the T2* weighted EPI sequence of time-resolved BOLD imaging was 2000 ms. The acquisition time was of ∼7 minutes. Data from all MRI modalities was preprocessed using FSL and QA followed Smith et al. (Smith et al., 2004). Subjects were removed if any of their scan acquisitions contained excessive artifacts including slice dropouts on the diffusion-images (defined by zebra-like blurring or complete dropout; Pannek et al., 2012), the presence of orbitofrontal EPI signal dropout (Weiskopf et al., 2007), excessive motion on T1-images (i.e., ringing), or severe geometric warping. For details of additional fMRI preprocessing details (slice-timing correction, realignment and co-registration, linear detrending, head motion regression, probabilistic segmentation, spatial smoothing, etc.) please refer to Perry et al. (2017).

### Network parcellation

For structural and functional parcellation the AAL atlas was used focused on 16 limbic regions (see Fig. 6D) associated with early degeneration in AD according to Braak and Braak staging as we did before (Joelle Zimmermann et al., 2018). The regions of interest included: Cingulate cortices (anterior, medial and posterior), Parahippocampal gyrus (including Entorhinal cortex), Hippocampus proper, amygdala, and temporal pole (superior and middle). In this study, as pertinent given the spatial organization retrieved in many of the analysis results, we categorize regions as belonging: either to *“Zone I”*, including ventral regions (superior and medial portion of the temporal pole, parahippocampal gyrus, hippocampus proper and amygdala in both hemispheres); or to *“Zone II”*, which included the six cingulate cortical regions (posterior, medial, and anterior) in both hemispheres; Fig. 6D). This subdivision in two separate zones allowed us the categorization of network links from dimers to higher-order arrangements (trimers, tetramers) determining “within zone” or “between zone” interactions based on the relative zone membership of the different nodes involved. We remark that the delimitations of Zone I and Zone II are inspired from data-driven considerations (the spatial organization of FC state centroids in Figs 2 and MC modules in Fig. 4) rather than from a-priori subdivisions.

### State-based dynamic Functional Connectivity

In this study, we applied two complementary dynamic functional connectivity (dFC) approaches to investigate non-stationarity of BOLD signals and capture the recurring, time-varying, functional patterns. The first one was the so called point-based method (PBM) introduced by Thompson and Fransson (2016), referred here as state-based dFC. This method assumes the existence of a small set of possible discrete FC configurations.

In this approach, BOLD signals of each subject were concatenated along the temporal dimension and transformed to z-scores using Fisher’s z-transformation to stabilize variance prior to further analysis. Following Thompson and Fransson (2016), we applied a *k*-means clustering algorithm on the concatenated time-series (Lloyd, 1982), to determine states based on global activity patterns (best partition out of 100 repetitions, max iterations 100). The optimal number of 4 clusters (*k* = 4) was validated based on detecting an elbow in the variation of the distortion score as a function of changing number of clusters *k* (Fig. S1B). Based on the collections of activity patterns at times assigned to each of the states, we computed Pearson correlation matrices, yielding *k* state-specific FC matrix FC^(λ)^ (λ =1…4). A state was hence characterized by the centroid activation pattern of time-frames within the state cluster and by its state-specific FC matrix (see Fig. 2A and Fig. S1A). To characterize the spatial properties of state-specific FC, we then used a graph-theoretical approach and measured global and local efficiencies (Achard and Bullmore, 2007; Latora and Marchiori, 2001) of the four FC^(λ)^ networks (Fig. 2B) using the Brain Connectivity Toolbox (Rubinov and Sporns, 2010).

To study the properties of the sequence of the dynamical states and the resulting temporal network dynamics, we followed Thompson and Fransson (2016) to construct a temporal network by using as network frame at a time *t* the graph FC^(λ)^ of the state λ observed at time *t.* This procedure transformed each fMRI session with *T* timestamps into a temporal network with *T* frames, each including *l* = *n*(*n* – 1)/2 links between each undirected pair of nodes. These temporal networks were binarized thresholding links as a function of an arbitrary common threshold *θ*. We then computed various temporal metrics describing network dynamics. First, we calculated the *mean dwell-time* for each subject by averaging the number of consecutive time-points belonging to a given state before changing to a different state (Fig. 2C). Second, we computed the proportion of time spent in each state as measured by percentage relative time (*state census*) (Fig. 2D). Third (for this step, binarization was necessary), we measured inter-contact times (ICT) of different links. ICTs for each link was defined as the temporal distance between events of link activation (i.e. link strength going above threshold) and offset (link strength going below threshold). For each link and each value of threshold *θ*, we computed the *mean activation* μ as a measure of mean duration of a link’s active intervals; the *total active time fraction* τ which is the total fraction of time in which a link was active relative to the duration of the imaging acquisition; and the *burstiness coefficient* (Goh and Barabási, 2008) assessed by:

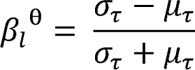

where *σ_τ_* and *µ_τ_* are, respectively, standard deviation and the mean of the ICTs along the considered temporal network instance. The burstiness coefficient is bounded in the range −1 ≤ *β* ≤ 1, such that *β* = −1 indicates a periodic/tonic link activation time-course, *β* = 0 a sequence with Poisson-like activation, and *β* = 1 corresponds to bursty (time-clustered) events of link activation (Fig. 3C). We finally evaluated also the memory coefficient (see always Goh and Barabási, 2008), which is the autocorrelation of the sequence of link activation times; i.e., if *E^(l)^_s_* is the duration of the *s-*th individual activation of link *l*, then memory coefficient for link *l* is λ*^(l)^* = CC(*E^(l)^_s_, E^(l)^_s+1_*), where CC denotes normalized Pearson correlation. Analogously, the burstiness and memory coefficients were averaged across links (or link classes, such as between-zone or within-zone links).

Unlike the mean dwell-time or state census, mean ICTs and the quantifications computed from them, depend on the specific choice of threshold *θ*. In absence of clear criteria to choose an optimum threshold value, we varied systematically *θ* in the range 1% *MAX* < *θ* < 10% *MAX* and *MAX* is the global maximum FC entry across the retained FC^(λ)^ state. The maximum value was equal to MAX = 0.87, therefore the range was 0.0087 < *θ* < 0.087. Absolute values of μ, τ and *β* varied with *θ*, however we pooled them together across different threshold values by computing relative variations (at each fixed *θ*) with respect to reference values (threshold-dependent), based on the NC group. For instance, for burstiness, we computed the relative excess burstiness for SNC, aMCI, and AD groups with respect to NCs (Fig. 2E) as:

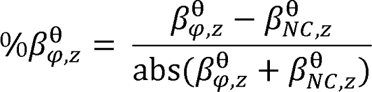

where *φ* = SNC, aMCI, AD and *z* refer to intra-zone, and subsets of inter-zone links. Analogously, we evaluated excess deviations for the SNC, aMCI, and AD relative to the NCs, across all possible thresholds, for μ and τ.

### State-free dynamic Functional Connectivity

In a second approach, we assumed that FC networks are continually morphing in time, without priors on the existence of discrete state switching events, following Battaglia *et al*. (2020), that conceptualized the evolution of FC as a stochastic walk in the high-dimensional space of possible network configurations. This stochastic walk however is not trivial, as different inter-regional links covary according to a specific higher-order correlation structure called *meta-connectivity* (Lombardo et al., 2020). State-free and smoothly varying dFC temporal networks were extracted using a sliding window approach, adopting the random-walks and meta-connectivity approaches (Battaglia et al., 2020; Lombardo et al., 2020; Petkoski et al., 2023) released within the dFCwalk toolbox (Arbabyazd et al., 2020).

A short window of size *ω* = 5 TRs (10 s) was stepped without overlap over the BOLD time-series acquired in each fMRI session and then functional connectivity matrices (FC) were computed as window-restricted Pearson’s correlation matrices between BOLD time-series segments. Each temporal frame provides hence *l* = *n*(*n* – 1)/2 undirected time-resolved link estimates, which can be collected into a *1* × *T* dFC stream, where *T* is the total number of windows. Each row of this stream provides the time-series of smoothed “instantaneous” variation of each FC link and the covariance between these variations can be described by a *l* x *l* matrix called the meta-connectivity (MC, Fig. 3B, (Lombardo et al., 2020)). The general entry of MC is given by:

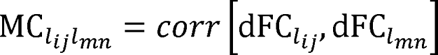

for every pair of links (*l_ij_* and *l_mn_*) formed respectively between the regions *(ij)* and *(mn).* Our choice of window length *ω* = 5 TRs was guided by: first, the observation from state-based dFC analyses that ∼90% of epochs within a state lasted less than 5 TRs (Fig. S2A), so that fast dFC dynamics may be lost using much longer windows; second, one-way ANOVA analysis on MC for a range of windows (from 3 to 20 TRs) showed that the best discrimination between SNC, NC, MCI and AD groups was achieved for *ω* = 5 TRs, with high between-group standard deviation and low within-group standard deviation (Fig. S2B). These analyses together suggest a small window of size *ω* = 5 TRs is both needed and sufficient to describe ongoing fast dFC fluctuations.

Following and based on the correlation matrix between “dimers” (dynamic FC links between two regions *i* and *j*), the entries MC_ij, kl_ of the MC matrix are either computed based on the dynamics of four regions involved in the links *(ij)* and *(kl)*, or at least three regions, when the two considered dimers share a common vertex (e.g*. i = k*). MC can thus be seen as a collation of higher-order interactions within the system, involving more than “two parts” (tetramers or trimers). In the case of a trimer, the region on which the two dimers converge to a “root” region, and the other two regions are the “leaves” of the trimer. In the case of a tetramer, each of the two non-incident dimers are called a “base”.

### MC modularity

We used a graph-theory approach to quantify the communities of MC matrices. MC for all subjects were constructed and then averaged for each of the four subject’s groups (Fig. 3B). To detect the modular structures of MC, we used the community Louvain algorithm (Rubinov and Sporns, 2011). We used a parameter Γ = 1.4, determined heuristically to yield a modular partition naturally interpretable in anatomical terms. To quantify the modularity changes across the groups, we computed the index of modularity (Q*) as measure of degree of intra-module connectivity. Since MC is a signed matrix, we applied disproportionate scaling to the positive and negative values of modularity indices to consider a lower contribution of negative meta-link weights to the index of modularity (Rubinov and Sporns, 2011). To quantify the degree of inter-modular connectivity of group averaged MCs, we computed the Participation coefficient of each dFC dimer node following (Guimera, Roger; Amaral et al., 2005). This metric can be computed exactly as for an ordinary graph keeping in mind that FC links and meta-links among them are, respectively nodes and links in the MC graph. The Participation coefficient is close to one when meta-links of a link are distributed uniformly, therefore, integrated across MC modules and it is zero when all the meta-links of a link are segregated within its own MC module.

### Meta-strengths

MC describes largely delocalized interactions but, for enhanced interpretability, it is important to describe the overall contribution of individual regions to the different higher-order interactions. Hence, we defined various indices of meta-strength.

Concerning trimer interaction, a natural definition of the trimer strength of a region *j* is given by:

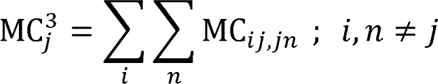

Here *j* is the root of the summed trimers, hence the name of “root-pinned” trimer strength (to contrast it with alternative definitions, not used in this study, where the pinned region may lie at a leaf). Analogously, we can define tetramer strengths of a link *(ij)*:

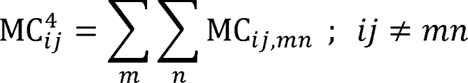

denoted as “base-pinned” as the frozen link is a dimer base of the tetramer.

A trimer is defined between zones or within zones depending on the zones to which its leaves belong. If all leaves are in the same zone (independently from where the root is) then the trimer is considered within zone, otherwise it is considered between zones. For tetramers we distinguished tetramers with base within a zone (if both bases are within zone dimers) or base between zones (if both bases are between zones). There are more combinatorial cases for tetramers that were ignored in this study for simplicity.

### Comparing MC and FC

We also computed more conventional FC strengths (dimer strengths) for each node as:

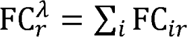

Where *λ* is an index referring to intra-zone if *i*and *r* are in the same zones (Fig. S4A), or inter-zone if they belong to different zones (Fig. 5A). To evaluate MC-FC redundancy on the single subject-level, we computed the Pearson’s correlation between roots-pinned trimers and FC node-degrees for all nodes and subject (Fig. 4A)., by the following formula:

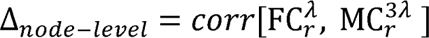

For the tetramers case, the same MC-FC comparison was done for edges computing:

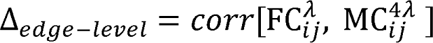

on the subject-level and for two intra- and inter-zone subsets (Fig. S3A).

We also introduced notions of genuine trimer and tetramers, to identify higher-order interactions that were not completely explained by existing motifs of dimer interactions. We separated trimers into three groups: 1) within zone, 2) leaves in same zone, and 3) leaves in two zones. For a given trimer with *r* as root and *i, j* as leave regions, we defined the following condition:

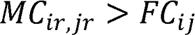

for a trimer to be considered “*genuine*”, meaning that the trimer interaction coupling *i* and *j* via *r* is not a mere byproduct of the dimer between *i* and *j* but it is actually stronger (another interpretation is that the interaction path between *i* and *j* is “shorter” when the interaction is mediated by *r* than when it is direct). Analogously, we separated tetramers into two groups: 1) base in two zones, and 2) base in same zone. For a give tetramer with (*i*, *j*) and (*m*,*n*) dimers, we the defined the following genuinity condition:

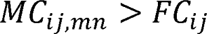

## Data Availability

All data produced in the present study are available upon reasonable request to the authors

## Supplementary tables

**Table S1.** Differential impact of pathology on FC dimers and MC trimers and tetramers.

|  | Intra-zone |  |  |  |
| --- | --- | --- | --- | --- |
|  | SNC | NC | aMCI | AD |
| FC | 0.543±0.170 | 0.564±0.155 | 0.549±0.186 | 0.490±0.180 |
| Trimers | 0.359±0.139 | 0.348±0.126 | 0.333±0.146 | 0.318±0.144 |
| Tetramers*** | 0.222±0.096 | 0.196±0.087 | 0.186±0.077 | 0.156±0.088 |
|  | Inter-zone |  |  |  |
| FC** | 0.101±0.114 | 0.083±0.135 | 0.054±0.126 | 0.021±0.088 |
| Trimers** | 0.039±0.078 | 0.019±0.083 | 0.013±0.072 | -0.012±0.052 |
| Tetramers*** | 0.183±0.134 | 0.187±0.117 | 0.138±0.137 | 0.139±0.120 |

*Average strengths of dimer, trimer and tetramer interactions, by clinical group and relation to anatomical zones. Values are means ± SD; \* significantly inter-group variations with $P < 0.05$ ; \*\* with $P < 0.01$ ; \*\*\* with $P < 0.001$ (one-way ANOVA test).*

**Table S2.** Memory coefficients for dynamic links in the four groups.

|  | Intra-zone |  |  |  |
| --- | --- | --- | --- | --- |
|  | SNC | NC | aMCI | AD |
| 5% | 0.1561 | 0.1310 | 0.1168 | 0.1037 |
| 50% | 0.1653 | 0.1383 | 0.1238 | 0.1098 |
| 95% | 0.1746 | 0.1457 | 0.1307 | 0.1158 |
|  | Inter-zone |  |  |  |
| 5% | 0.1407 | 0.1391 | 0.1404 | 0.0901 |
| 50% | 0.1452 | 0.1428 | 0.1443 | 0.0928 |
| 95% | 0.1498 | 0.1465 | 0.1481 | 0.0954 |

*The memory coefficient, by clinical group and relation to anatomical zones. Values are means and the confidence intervals; Intra-zone: SNC >>> NC, aMCI >>> NC, AD >>> NC ; Inter-zone : aMCI >>> NC, AD >>> NC ; where, >>> means p-value smaller than 0.001.*

## Supplementary figures

**Fig. S1.**
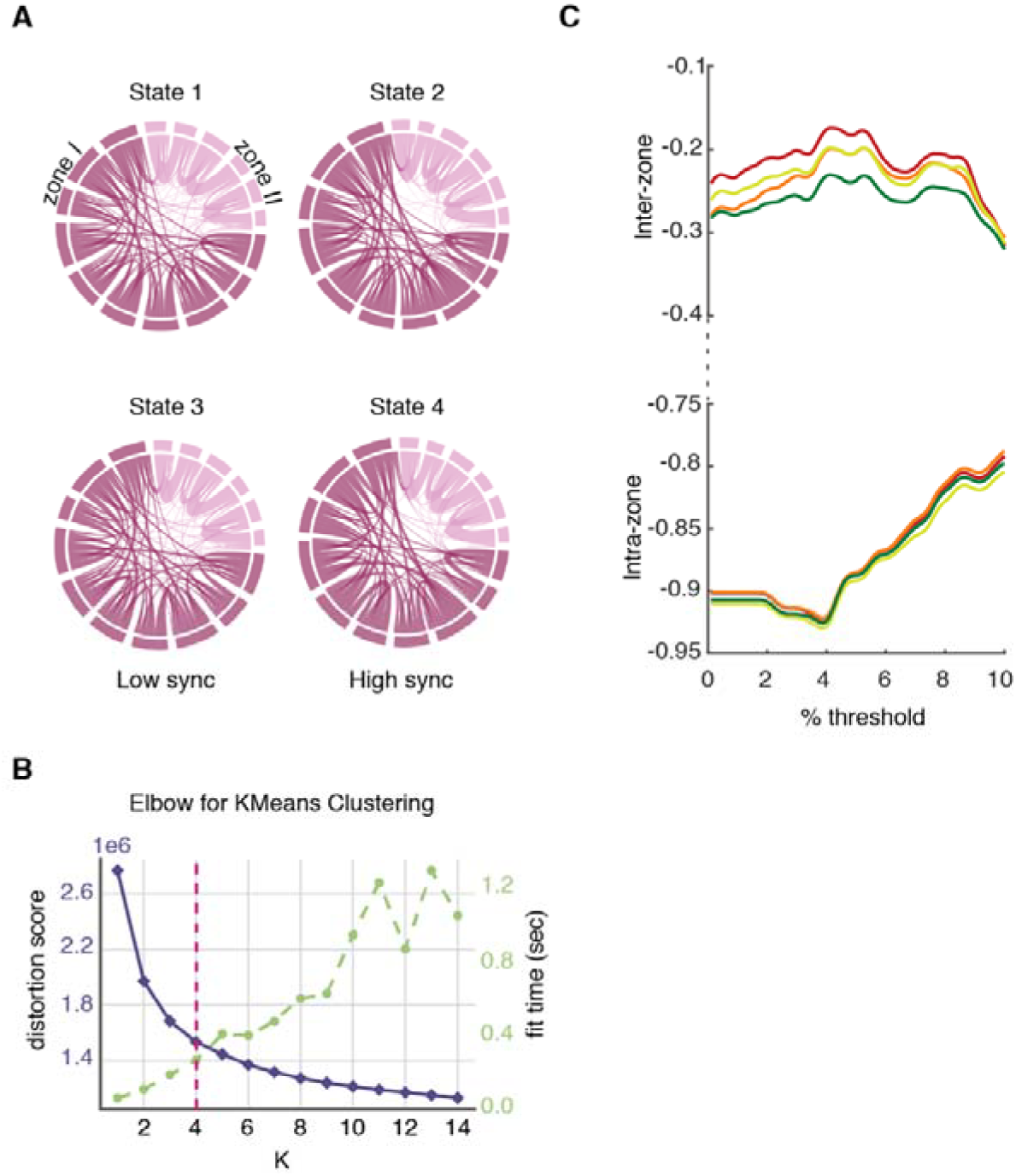
Addition information on state-based dFC analyses. **(A)** Chord diagrams of FC^(λ)^ states as an alternative illustration of Fig. 1A. Dark pink regions correspond to Zone I and light pink regions to Zone II. States 1 and 3 with low synchronization have stronger inter-zone connections than states 2 and 4 with high synchronization. **(B)** We used an elbow criterion based on the Silhouette score to guess the optimal number of clusters. The distortion (linked to the distance between cluster centroids) slows down its decrease with *k* while the time of clustering keeps growing, leadings to estimate a number of retained clusters around four (**C**). We show here the dependence of the average burstiness coefficient β for all groups on different choices of binarization thresholds θ. which were averaged over dFC dimers into two intra- and inter-zone categories of links is shown (colored solid lines; green: SNC, yellow: NC, orange: aMCI, red: AD). The fact that the gap and the relative ranking between curves for the different groups remain consistent over different thresholds justifies the use of relative excess values for the analyses of Figure 3E.

**Fig. S2.**
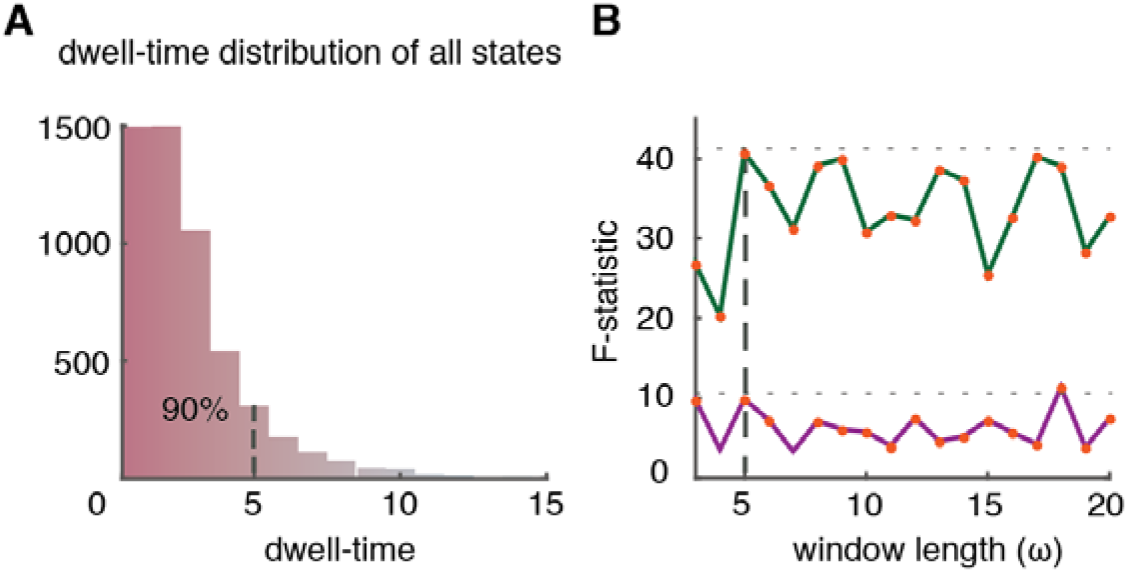
Length of window in MC approach. (**A**) Distribution of the duration of mean dwell-times in a consistent state (from the state-based PBM method), pooled over subjects and states (see Fig 2C). We see that ∼90% of epochs last less than 5 TRs. (**B**) We applied one-way ANOVA on average MC strengths to determine the existence of inter-group differences. Shown here is the value of the F-statistic for existence of inter-group differences, as a function of changing window size, from 3TRs to 20TRs. We performed the analysis separately for *intra-zone* (green line) and *inter-zone* (violet line) subsets of trimers. Using larger windows would not improve the statistical detection of inter-group differences. A short window of length ω = 5TRs is thus already sufficient to capture between-group differences, maintaining at the same time the capability to track the very fast dFC fluctuations revealed by Fig. S2A.

**Fig. S3.**
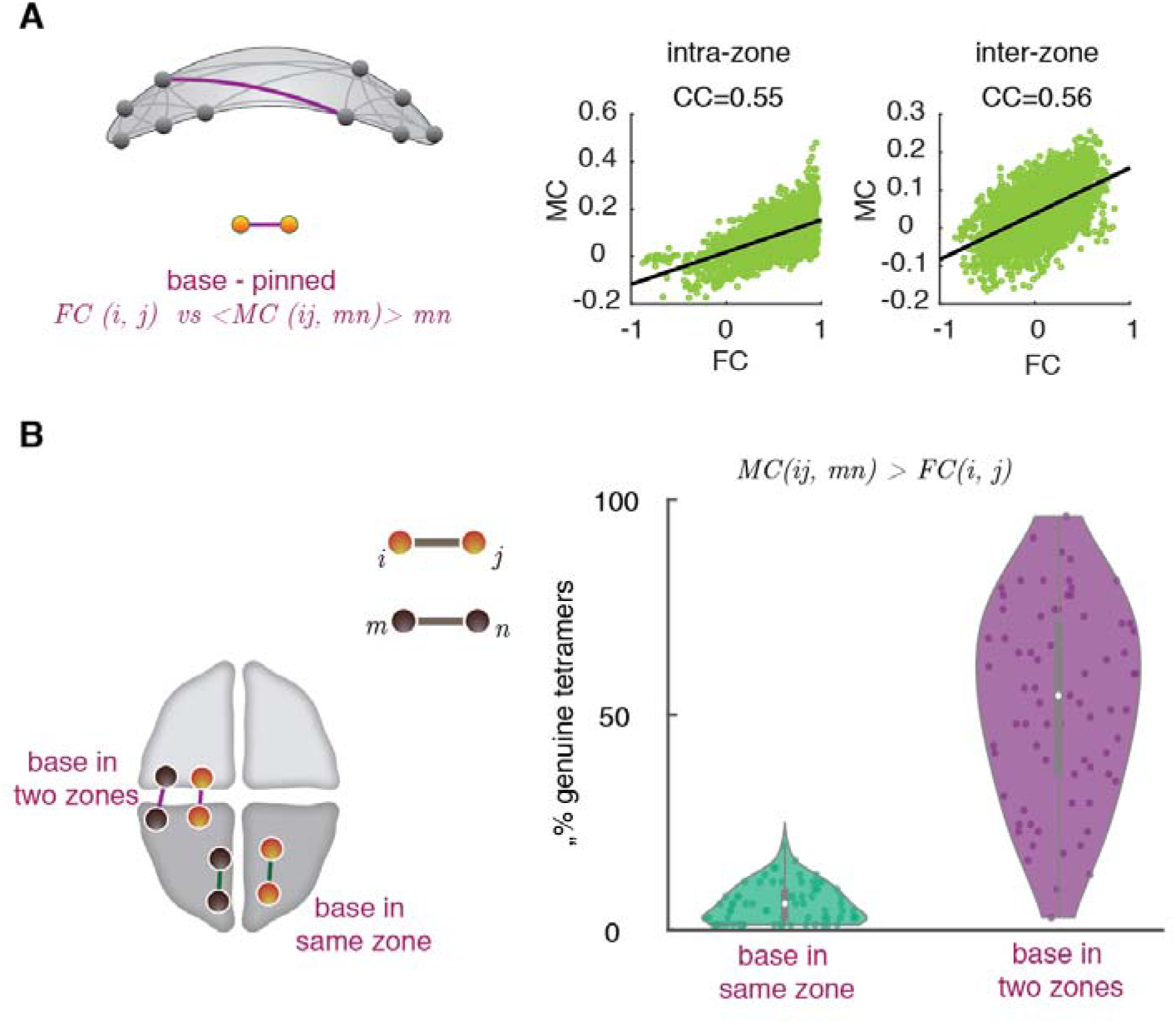
State-free dFC: Inter-relations between dFC tetramers and FC dimers. (**A**) Similarly to the MC-FC comparison at the trimer level (see Fig. 5A), we compared dimer and tetramer strengths now for edges. The scatter plots show values of FC dimers paired with the corresponding base-pinned tetramer strength of that dimer (i.e. the overall meta-coupling of that dimer to other remote and non-incident dimers). Again, values are separated for intra- and inter-zone dimers and tetramers. Unlike for trimers, strong dimers are also the ones with the strongest tetramer strengths, as revealed by significant positive correlations. (**B**) Generalizing Fig. 5B for trimers, we also computed the fraction of genuine tetramers. The *base in same zone* subset of tetramers contained a low fraction of genuine tetramers, while this fraction raised for tetramers with an inter-zone base.

**Fig. S4.**
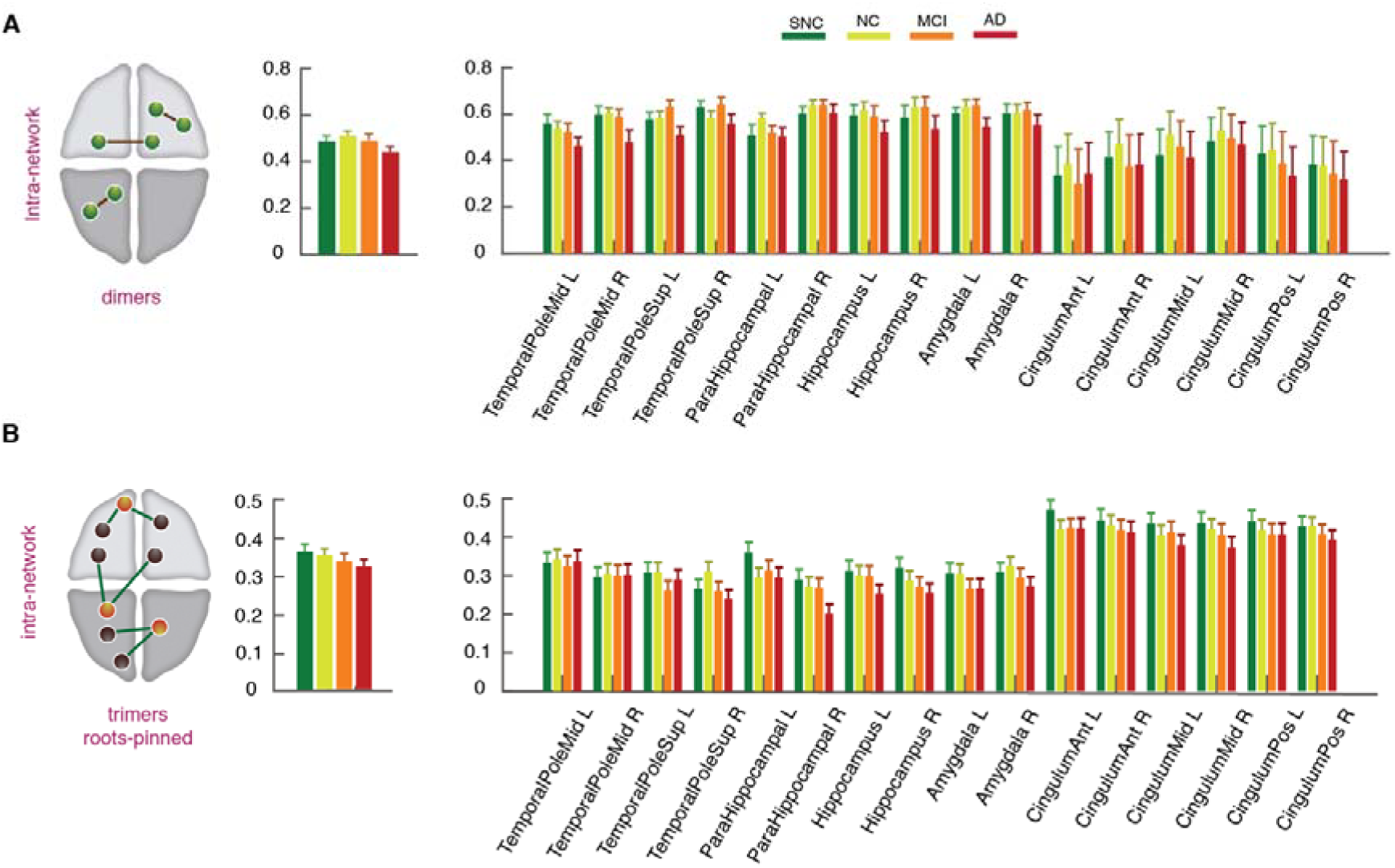
State-free dFC: intra-zone FC dimers and dFC trimers strengths. (**A**) and (**B**) The FC dimers and dFC trimers for the intra-zone subset did not show any significant reduction of strength from SNC-to-AD group, despite moderately decreasing average values, both globally (left) and locally at the single region level (right).

